# Longitudinal progression of digital arm swing measures during free-living gait in early Parkinson’s disease

**DOI:** 10.64898/2026.01.06.26343500

**Authors:** Erik Post, Twan van Laarhoven, Nienke A. Timmermans, Yordan P. Raykov, Max A. Little, Jorik Nonnekes, King Chung Ho, Tom M. Heskes, Bastiaan R. Bloem, Luc J.W. Evers

## Abstract

Asymmetric arm swing reduction is a hallmark of early-stage Parkinson’s disease (PD). We evaluated wrist sensor-based arm swing measures during free-living gait as digital progression biomarkers in 623 early-stage PD participants and 50 controls monitored continuously for one year (controls) or two years (PD). Biweekly measures were extracted from gait segments without other arm activities. Arm swing measures were reduced in PD, reduced on the most affected side, moderately correlated with clinical motor severity, and highly reliable. In unmedicated participants, most affected side measures declined over one year (standardized response means: −0.32 to −0.78) and two years (−0.51 to −1.10), exceeding an MDS-UPDRS Part III subscore at one year (−0.21) and matching it at two years (−0.83). In medicated participants, arm swing increased on the most affected side, which was associated with higher dopaminergic doses and dyskinesia. Overall, free-living arm swing measures are promising progression biomarkers in early, untreated PD.

## Introduction

Parkinson’s disease (PD) is the second most common and fastest-growing neurodegenerative disease globally [1, 2], causing profound and progressive disruption to the lives of those affected [3, 4]. Despite its growing burden, treatment remains symptomatic, highlighting the urgent need for disease-modifying therapies. Progress in this area depends not only on the development of novel interventions but also on the implementation of robust clinical trial designs. Crucially, the success of these efforts hinges on the availability of accurate, reliable measures sensitive to disease progression and treatment response [5].

Currently, disease severity and treatment efficacy are assessed primarily using clinical scales such as the Movement Disorder Society-sponsored Unified Parkinson’s Disease Rating Scale (MDS-UPDRS). However, these in-clinic, episodic assessments fail to capture the variable course of symptoms in daily life and suffer from observation effects [6–8]. In addition, their ordinal, subjective nature and high intra-subject variability [9] limit their reproducibility and sensitivity to subtle changes, raising concerns about their suitability for evaluating disease-modifying interventions, particularly as phase 2 and phase 3 trials continue to demonstrate neutral or negative results [10, 11].

To overcome these limitations, wearable sensor technologies have emerged as a promising complementary approach. They enable continuous data collection in real-world conditions, which can be used to derive high-resolution insights into motor and non-motor symptoms [7, 12]. In addition, digital outcomes may be more sensitive to progression in early PD, potentially reducing the sample sizes required for clinical trials [13]. They may also enable earlier detection of disease onset, creating new opportunities for prevention trials [7, 13]. And, importantly, because they can be deployed in free-living conditions, digital measures can capture aspects of daily living meaningful to patients [3, 14].

AAmong the motor symptoms that digital measures can capture, reduced arm swing during gait stands out as a promising target for multiple reasons. It is one of the earliest signs of PD, typically appearing up to seven years prior to diagnosis [15–17]. Reduction in arm swing is initially often unilateral, and although the contralateral side eventually also becomes affected, arm swing remains asymmetric for most individuals throughout the disease [18]. Subtle changes have even been observed in non-manifesting LRRK2-G2019S mutation carriers, further supporting a role for arm swing reduction as a potential early biomarker [19]. As the disease progresses, contralateral involvement emerges, making arm swing a useful marker of lateralized progression [20, 21]. Furthermore, arm swing is responsive to dopaminergic treatment, underscoring its sensitivity to change [22, 23]. Analyses of clinic-based wearable sensor data show that arm swing may capture these disease-related changes more robustly than conventional MDS-UPDRS gait items [24]. Importantly, reduced arm swing compromises gait stability and increases fall risk [25], which are leading causes of injury in PD [26].

Although arm swing can be evaluated in the clinic, studies have demonstrated discrepancies between in-clinic and at-home gait assessments [6, 24, 27, 28], reinforcing the need for continuous, real-world monitoring. Wrist-worn sensors provide a user-friendly solution for continuous, long-term monitoring of arm swing in daily life [29]. For example, both the Parkinson’s Progression Markers Initiative (PPMI) [30] and the Personalized Parkinson Project (PPP) [31] demonstrated excellent long-term compliance, with dropout rates below 5% and median wear times of 22 hours per day over two years in the PPP [29].

Yet, despite its clinical relevance and feasibility for digital tracking, continuous measures of arm swing in free-living conditions remain underexplored, and no longitudinal studies to date have assessed their reliability or sensitivity to progression [13]. A key challenge in realizing longitudinal arm swing assessments is the need for validated algorithms that can accurately quantify arm swing in free-living conditions. Unlike structured clinical assessments, free-living gait is interspersed with diverse arm activities that can obscure true arm swing patterns. To address this, we previously developed and validated a signal processing pipeline for wrist-worn inertial measurement unit (IMU) data collected in free-living settings [22]. This pipeline automatically detects gait segments, filters out periods with other arm activities, and computes the range of motion (RoM) of arm swing.

The present study evaluates the ability of digital arm swing measures, collected during free-living gait, to quantify disease progression. We use data from the PPP, in which 623 early-stage PD participants wore a wrist device continuously for at least two years under free-living conditions, alongside 50 controls monitored for one year. First, we assess construct validity of the digital measures by examining differences between PD and controls, between the most and least affected side, and by determining the correlation with clinician- and patient-reported outcome measures. We then assess test-retest reliability by calculating week-to-week intra-subject variability. Finally, we assess sensitivity to disease progression by estimating longitudinal changes in unmedicated and medicated participants, and evaluate the impact of dopaminergic medication adjustments on the digital measures in medicated participants.

## Methods

### Data

We used data from three prospective longitudinal cohort studies. The first is the Personalized Parkinson Project (PPP), a cohort of 520 individuals with early-stage PD (≤ 5 years since diagnosis), including 25 participants who had not initiated dopaminergic medication at baseline [31]. To study longitudinal changes in arm swing without the effects of dopaminergic treatment, we additionally included data from the PPP De Novo cohort comprising 103 recently diagnosed (≤ 2 years since diagnosis), treatment-naïve PD participants [32]. During recruitment, individuals expected to start treatment within 52 weeks from baseline were excluded [32]. Finally, to assess the specificity of observed changes to PD, we included data of 50 control participants from the PPP PSP cohort [33], where individuals with neurological or musculoskeletal disorders or other conditions that impair movement were excluded, as determined by the investigator. Controls for the PPP PSP cohort were often partners from participating individuals with progressive supranuclear palsy (PSP).

Individuals with PD wore the Verily Study Watch on their preferred wrist for at least two years, with instructions to wear it continuously and charge it for up to two hours during the day. The control group followed the same protocol for one year. Adherence was high in the PPP study: only 4% of participants dropped out, and those completing the two-year period had a median wear time of 22.0 hours per day [29]. In-clinic assessments were conducted annually, starting at baseline, and included the MDS-UPDRS and the Hoehn and Yahr (H&Y) staging scale for PD participants (Figure 1).

**Figure 1:**
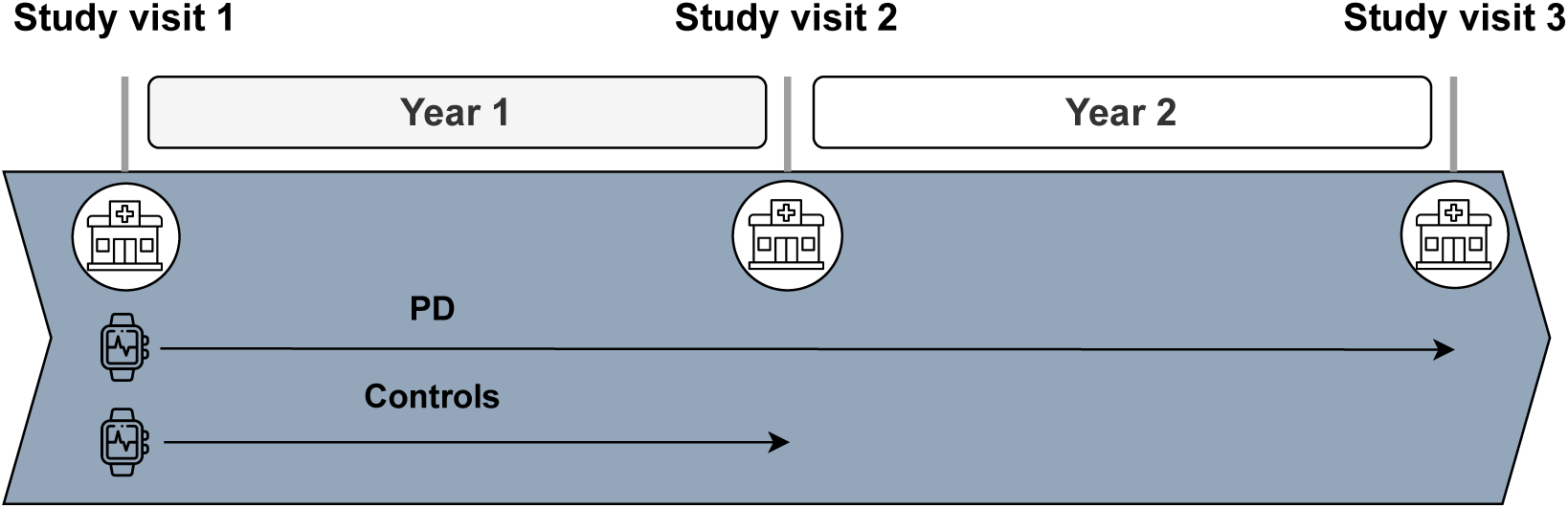
Study design overview. At baseline (study visit 1), participants with PD underwent in-clinic assessments and received a smartwatch with instructions to wear it continuously for at least two years. During follow-up, in-clinic assessments were repeated annually (study visits 2 and 3). Control participants wore the watch continuously for one year and underwent in-clinic assessments at baseline (study visit 1) and after one year (study visit 2).

#### Digital arm swing measures

We used triaxial accelerometer and gyroscope data sampled at 100 Hz (range of ±4g, ±1000 deg/s). Arm swing range of motion (RoM) was computed using the ParaDigMa toolbox [34], which quantifies arm swing by detecting gait segments in free-living conditions from wrist IMU data. The toolbox includes a method for identifying and excluding gait segments with other arm activities (e.g., hands in pockets, carrying an object) that are highly prevalent in free-living conditions and can confound arm swing measurements. Arm swing RoM was calculated using a previously validated method based on absolute peak-to-trough angular displacements in the arm swing direction [35]. The full signal processing pipeline and its validation during unscripted activities in the home environment are described in detail elsewhere [22]. The generalizability of the gait detection algorithm to the current dataset is demonstrated in Supplementary Figure 1 and Supplementary Table 1.

To manage computational load and data volume, we quantified arm swing every other week, except at baseline, where two consecutive weeks were analyzed to assess test-retest reliability. For each participant, we derived 6 digital measures by stratifying arm swing RoM along two dimensions:

1. Aggregation method: we aggregated across one week to estimate the typical (median) arm swing, arm swing capacity (95^th^ percentile), and typical (median) swing-to-swing variability (coefficient of variation (CoV)). To obtain the swing-to-swing CoV, we divided the standard deviation by the mean range of motion per gait segment, and computed the median across gait segments.
2. Gait segment duration: we aggregated for short (*<* 20 s) and long gait segments (≥ 20 s) separately, because prior studies suggest that PD-related gait impairments may vary between short and long gait segments in free-living settings [27].

To assess the impact of watch side, we analyzed participants wearing the watch on the most and least affected side separately, which we determined using the unilateral MDS-UPDRS Part III subscore in the OFF state at baseline (see section *Clinician- and patient-reported measures* for a definition of this clinical score). In case of equal lateral scores, subjective participant reports were used to resolve side assignment; participants without perceived and measured asymmetry were excluded from the analyses.

#### Clinician- and patient-reported measures

To provide a clinical reference for the impairments targeted by the digital measures, we calculated a unilateral subscore from MDS-UPDRS Part III bradykinesia and rigidity items (see Supplementary Table 2 for a full list of items comprising the subscore). This subscore, reflecting bradykinesia and rigidity in both upper and lower limbs, was computed separately for the OFF state (for medicated participants at least 12 hours after last dopaminergic medication intake) and the ON state (as reported by the participant following medication intake; only for medicated participants). To assess how digital arm swing measures relate to experienced symptoms, we also examined their correlations with the sum of items related to bradykinesia and rigidity from the MDS-UPDRS Part II (items 2.4–2.7) and the PDQ-39 (items 11–16).

### Cross-sectional analyses

The cross-sectional analyses were based on digital measures obtained from the second week of follow-up, to exclude potential effects from the baseline study visit. For test-retest reliability, we additionally included the third week of follow-up.

#### Study sample

Participants were excluded from the cross-sectional analyses based on the following criteria:

- Use of a walking aid (MDS-UPDRS Part III item 2.12 ≥ 2) or presence of significant dyskinesia at baseline (MDS-UPDRS Part IV item 4.1 ≥ 2), because both factors likely interfere with accurate estimation of the arm swing range of motion.
- Unknown watch side.
- Insufficient sensor data: fewer than 3 days with ≥ 10 hours of wear time between 08:00 and 22:00 during either the second or third week [36].
- Insufficient filtered gait: fewer than 2 days with ≥ 2 minutes of gait without other arm activities during either the second or third week.

Participants with slight dyskinesia were retained, to align with the inclusion criteria of the ParaDigMa toolbox, to increase sample size, and to study how slight dyskinesia impacts our ability to measure disease progression. The thresholds for data sufficiency were chosen to ensure that weekly aggregates reflected both intra-day and inter-day variability, and to comply with quality criteria established by the ParaDigMa toolbox [34].

#### Construct validity

We evaluated whether the digital arm swing measures captured PD-related motor impairments. We compared measures between participants with PD wearing the watch on the most affected side, those wearing it on the least affected side, and controls. Digital measures were adjusted for covariates with significant between-group differences by including them as covariates in an ordinary least squares (OLS) regression model. Between-group differences in digital measures were quantified from model-derived estimates, and statistical significance was assessed with *t* -tests. For each measure, we additionally computed Spearman’s rank correlations with the unilateral MDS-UPDRS Part III subscore in the OFF and ON medication states and with the MDS-UPDRS Part II subscore.

#### Test-retest reliability

To assess week-to-week stability of the digital measures, we calculated the intra-class correlation coefficient (ICC) using a two-way ANOVA mixed effects model applied to weekly digital measures from the second and third week of follow-up [37].

### Longitudinal analyses

We evaluated the sensitivity to change of the digital arm swing measures over one and two years, separately for patients who were unmedicated and medicated during the full follow-up period. First, we applied an *ℓ*_1_ trend filter to the biweekly digital measures to extract each subject’s long-term trends. Based on these trends, we computed the one-year and two-year standardized response mean (SRM) to quantify sensitivity to change. In addition, we fitted a regression model to examine how disease characteristics and medication influence within-subject changes in the digital measures. In all longitudinal analyses, we used inverse probability weighting to adjust for potential bias due to censoring (e.g., due to initiation of dopaminergic medication in the unmedicated group, and developing dyskinesia or start using a walking aid in the medicated group).

#### Study sample

Participants were categorized into three groups: (1) participants who did not initiate dopaminergic treatment during the entire one- or two-year follow-up period (unmedicated group), (2) participants who had already initiated dopaminergic medication at baseline (medicated group), and (3) participants who initiated dopaminergic medication during the study. Participants in the third group were not included in the progression analyses because the initiation of dopaminergic has a large impact on PD motor symptoms (also known as the “honeymoon phase”), which complicates the detection of disease progression-related changes [38]. Additional exclusion criteria included:

- Use of a walking aid (MDS-UPDRS Part III item 2.12 ≥ 2) or presence of significant dyskinesia (MDS-UPDRS Part IV item 4.1 ≥ 2) at any time during the study.
- Diagnosis changed from PD to another disorder during follow-up.
- Unknown watch side, or a changed watch side during follow-up.
- Insufficient sensor data: fewer than 3 days with ≥ 10 hours of sensor data between 08:00 and 22:00 within each of the first three weeks (weeks 2, 4, and 6) or final three weeks (weeks 48, 50, and 52 for the one-year analysis; weeks 96, 98, and 100 for the two-year analysis).
- Insufficient filtered gait: fewer than 2 days with ≥ 2 minutes of gait without other arm activities within each of the first three weeks (weeks 2, 4, and 6) or final three weeks (weeks 48, 50, and 52 for the one-year analysis; weeks 96, 98, and 100 for the two-year analysis).

#### Deriving longitudinal trends

Weekly digital measures contain both long-term signal variability (e.g., progression of motor impairment, long-term changes in treatment) and short-term fluctuations (e.g., behavioral variability, short-term variations in motor impairment and measurement error). To isolate the long-term signal for each participant, we applied an *ℓ*_1_ trend filter [39], which estimates a piecewise linear trend by balancing two objectives: (1) minimizing the squared error between the estimated trend and observed data, and (2) encouraging smoothness by penalizing the number of changes in slope. This leads to the optimization problem

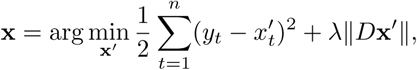

where **y** is the observed time series, **x** is the optimal estimated trend, *D* is a second-order difference operator, and *λ >* 0 is the regularization parameter.

We selected *λ* for each digital measure using cross-validation. A range of candidate *λ* values was evaluated (log10(*λ*) between 0 and 2). For each participant, we performed leave-one-time-point-out cross-validation: in each fold, a single observation was masked, the *ℓ*_1_ trend filter was fitted to the remaining observations, and the masked value was predicted using linear interpolation of the fitted trend. Prediction error was quantified as the absolute difference between the observed and predicted value and averaged across time points for each participant. Errors were then aggregated across participants, weighted by the number of available observations, and the *λ* minimizing the mean absolute error was selected. This procedure was performed separately for the medicated and unmedicated groups, resulting in a single *λ* per digital measure per group.

The optimal *λ* and corresponding error for each measure are shown in Supplementary Figure 2, and the proportion of variance explained by the fitted trends for each measure is reported in Supplementary Table 3. We inter- and extrapolated the fitted trends to weeks separated by at most two timepoints (e.g., extrapolating week 56 to weeks 58 and 60). All downstream analyses were based on the extracted trend signals.

#### Sensitivity to long-term changes

To quantify the sensitivity of digital measures to longitudinal change, we computed the SRM, defined as

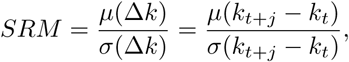

where *k_t_* and *k_t_*_+_*_j_* are digital measure values at weeks *t* and *t* + *j*, and *µ* and *σ* denote the group-level mean and standard deviation. Larger absolute SRMs indicate greater sensitivity to change.

SRMs were computed at one- and two-year follow-up using 5,000 bootstrap samples to estimate 95% confidence intervals (CIs). To compare sensitivity to change of digital measures with established clinical outcomes, SRMs were computed for the unilateral MDS-UPDRS Part III bradykinesia and rigidity subscores at one and two years. For the digital measures, we considered week 100 for two-year SRMs to maximize participant inclusion, and week 50 for one-year SRMs marking the halfway point. Clinical outcomes were included if assessments were conducted within 10 weeks of the one-year mark and within 20 weeks of the two-year mark. To assess wheter observed changes were specific to PD, one-year SRMs were computed for the control group.

#### Adjusting for censoring bias

In the unmedicated group, participants were censored at initiation of dopaminergic therapy (which is also common practice in clinical trials evaluating disease-modifying therapies [40]). Although this allowed us to study disease progression without the effects of medication, censoring could have introduced bias: individuals with faster motor progression likely have a higher likelihood to start dopaminergic treatment. Similarly, in the medicated group, participants were censored when they started using a walking aid or developed clinically significant dyskinesia, which also likely occur more often in participants with faster motor progression.

To account for potential bias introduced by censoring, we applied inverse-probability-of-censoring weighting (IPCW) [41]. IPCW reweights each participant’s contribution to the SRM at time *t* by the inverse of their estimated probability of remaining uncensored up to time *t*, conditional on set of time-dependent risk factors. This creates a pseudo-population that approximates the distribution that would have been observed in the absence of censoring. Because censoring mechanisms differed between groups, weights were estimated separately.

For the unmedicated group, we modeled time to censoring using a time-varying Cox proportional hazards model with both static and time-dependent covariates. The hazard function is defined as

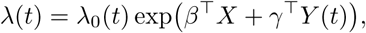

where *X* represents static covariates, *Y* (*t*) represents time-dependent covariates, and *λ*_0_(*t*) is the baseline hazard [42]. Based on the assumption that treatment initiation is likely influenced by current motor status, we included digital arm swing measures at time *t* and their recent change (from *t - 1* to *t* ). No static covariates were included.

In the medicated group, censoring occurred at a single timepoint, i.e. after the second study visit; if during the third visit, a participant was using a walking aid or demonstrated clinically significant dyskinesia, all data between the second and third visit were excluded. Here, because we predict a single point of censoring, we used logistic regression to model the probability of being uncensored using the same covariates as in the time-varying Cox proportional hazards model. Although the censoring mechanism is different, we postulate that both starting to use a walking aid and developing dyskinesia are the (indirect) result of more severe motor impairment.

Finally, the estimated probabilities of remaining uncensored up to time *t* were inverted and stabilized by multiplication with the corresponding marginal (unconditional) probability. Then, to calculate the corrected SRM between time *t* and *t + j*, we multiplied the stabilized weights at time *t + j* with the change in digital measures *k_t_*_+_*_j_* - *k_t_*.

#### Robustness analyses

To assess the effects of filtering gait (i.e., detecting and discarding gait segments with other arm activities) and IPCW reweighting on sensitivity to change, one- and two-year SRMs were compared between digital measures of filtered and unfiltered gait segments, and between weighted and unweighted measures.

#### Impact of dopaminergic medication on longitudinal change

To examine how dopaminergic medication influences the ability to measure progression in the medicated group, we fitted OLS regression models. The dependent variable was the two-year change in each digital measure. Predictors included change in levodopa equivalent daily dosage (LEDD) and the development of slight dyskinesia, in addition to time since diagnosis and watch side (most versus least affected side). We included interactions between watch side and change in LEDD, and between watch side and the development of slight dyskinesia.

### Code availability

All analyses were conducted using Python 3.12 and the scipy [43], statsmodels [44], and lifelines [45] packages in a Poetry-managed environment to ensure reproducibility. The arm swing quantification pipeline is part of the ParaDigMa toolbox [34]. Code for reproducing results (Python) and fitting the *ℓ*_1_ trend filter (MATLAB) is publicly available at https://github.com/AI-for-Parkinson-Lab/longitudinal_arm_swing.

## Results

### Cross-sectional

A total of 420 out of 623 participants with PD (264 most affected side, 156 least affected side) and 44 out of 50 controls were included (Supplementary Figure 3). Baseline characteristics of all groups are shown in Table 1. As expected, compared to participants wearing the watch on the least affected side, those wearing it on the most affected side had higher MDS-UPDRS Part III subscores on the watch side in both ON and OFF states (*p <* 0.001). In addition, controls had more short gait segments than both PD groups (*p <* 0.001).

**Table 1:** Demographics of the study population. PD MAS: participants with PD wearing the watch on the most affected side; PD LAS: participants with PD wearing the watch on the least affected side; N: number of participants; SD: standard deviation; N/A: not applicable; MDS-UPDRS: Movement Disorder Society-sponsored Unified Parkinson’s Disease Rating Scale. Filtered gait: gait without other arm activities.

|  | PD MAS<br>(N = 264) | PD LAS<br>(N = 156) | Controls<br>(N = 44) |
| --- | --- | --- | --- |
| Women (N (%)) | 98 (36.7) | 72 (45.5) | 28 (63.6) |
| Age (years, mean ± SD) | 61.7 ± 8.9 | 61.6 ± 8.4 | 69.0 ± 7.5 |
| Watch on dominant side (N (%)) | 70 (26.5) | 22 (14.1) | 8 (18.2) |
| **Hoehn & Yahr stage (N (%))** |  |  |  |
| I | 23 (8.7) | 11 (7.1) | N/A |
| II | 218 (82.6) | 131 (84.0) | N/A |
| III | 23 (8.7) | 14 (9.0) | N/A |
| **MDS-UPDRS (mean ± SD)** |  |  |  |
| Part I | 6.9 ± 3.6 | 7.1 ± 3.9 | N/A |
| Part II | 6.2 ± 4.9 | 5.9 ± 4.7 | N/A |
| Part III OFF | 24.4 ± 9.2 | 23.3 ± 9.3 | N/A |
| Part III ON | 21.8 ± 9.7 | 19.5 ± 9.2 | N/A |
| Part III subscore OFF (watch side) | 11.8 ± 3.9 | 7.8 ± 4.2 | N/A |
| Part III subscore ON (watch side) | 10.5 ± 4.2 | 7.0 ± 4.3 | N/A |
| **Amount of gait (mean ± SD hours/week)** |  |  |  |
| gait segments $< 20$ s | 4.2 ± 1.4 | 4.5 ± 1.5 | 5.3 ± 1.7 |
| gait segments $\geq 20$ s | 5.6 ± 2.9 | 5.5 ± 3.2 | 5.2 ± 3.0 |
| **Amount of filtered gait (mean ± SD hours/week)** |  |  |  |
| Short gait segments ($< 20$ s) | 0.8 ± 0.5 | 1.1 ± 0.5 | 1.3 ± 0.6 |
| Long gait segments ($\geq 20$ s) | 2.2 ± 1.7 | 2.7 ± 2.0 | 2.2 ± 1.4 |

In addition, compared to controls, the PD groups included a higher proportion of men (PD most affected side versus controls: *p* = 0.001; PD least affected side versus controls: *p* = 0.051) and were younger (*p <* 0.001). In the PD group, participants wearing the watch on the most affected side more often wore it on their dominant side (*p* = 0.004) and had slightly higher total MDS-UPDRS Part III ON scores (*p* = 0.028), had slightly less gait (less short gait segments *p* = 0.011, less short gait segments without other arm activities *p <* 0.001, and less long gait without other arm activities (*p* = 0.034)). Therefore, between-group comparisons were adjusted for age, gender, dominant-side wearing, and, within the PD groups, additionally for disease severity (MDS-UPDRS Part III ON score). We did not adjust for gait quantity as this was not expected to affect the outcome measures.

#### Construct validity

Digital arm swing measures were lower in participants with PD wearing the watch on the most affected side than in controls, and lower in those wearing the watch on the most affected side than those wearing it on the least affected side (Figure 2; unadjusted results in Supplementary Figure 4). In short gait segments, 95^th^ percentile and coefficient of variation (CoV) range of motion were also reduced in participants with PD wearing the watch on the least affected side compared with controls. In long gait segments, only the CoV range of motion was significantly reduced on the least affected side compared with controls.

**Figure 2:**
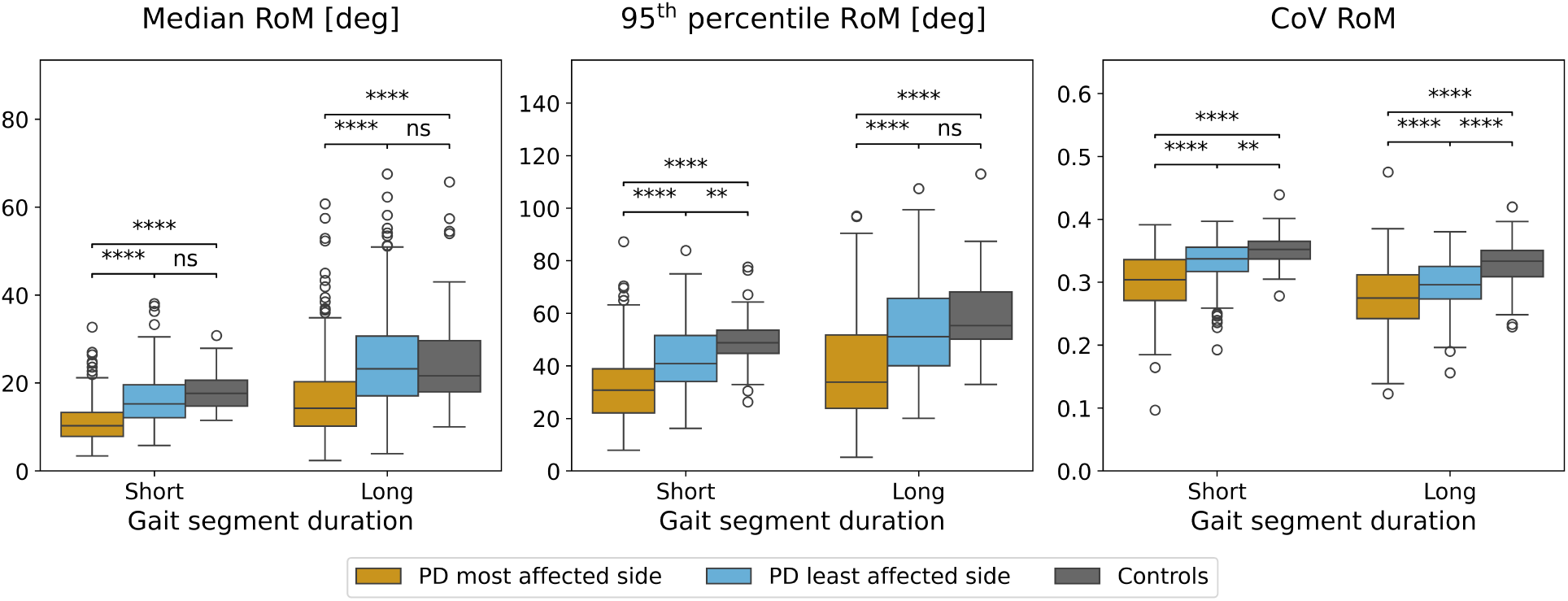
Between-group differences of digital measures. Differences were adjusted for age, gender, dominant side, and, within PD groups, MDS-UPDRS Part III ON scores using ordinary least squares regression. Comparisons were made using paired *t* -tests. Notation for two-sided thresholds of statistical significance: *p <* 0.05 (*), *p <* 0.01 (**), *p <* 0.001 (***), and *p <* 0.0001 (****): RoM: range of motion; CoV: coefficient of variation; deg: degrees; short: gait segments *<* 20 seconds; long: gait segments ≥ 20 seconds; ns: not significant.

In unmedicated PD, higher MDS-UPDRS Part III subscores on the watch side were associated with a smaller median, 95^th^ percentile and CoV RoM in short gait segments, and a smaller CoV RoM in long gait segments (Figure 3). Notably, higher MDS-UPDRS Part III subscores on the non-watch side were associated with larger median and 95th percentile RoM in participants wearing the watch on the least affected side, possibly indicating compensation effects. A similar pattern was observed for the MDS-UPDRS Part II and PDQ-39 subscores. In medicated PD, only the median and 95^th^ percentile RoM retained consistent associations, whereas variability was not significantly correlated with any of the clinical evaluations of bradykinesia and rigidity (Supplementary figure 5).

**Figure 3:**
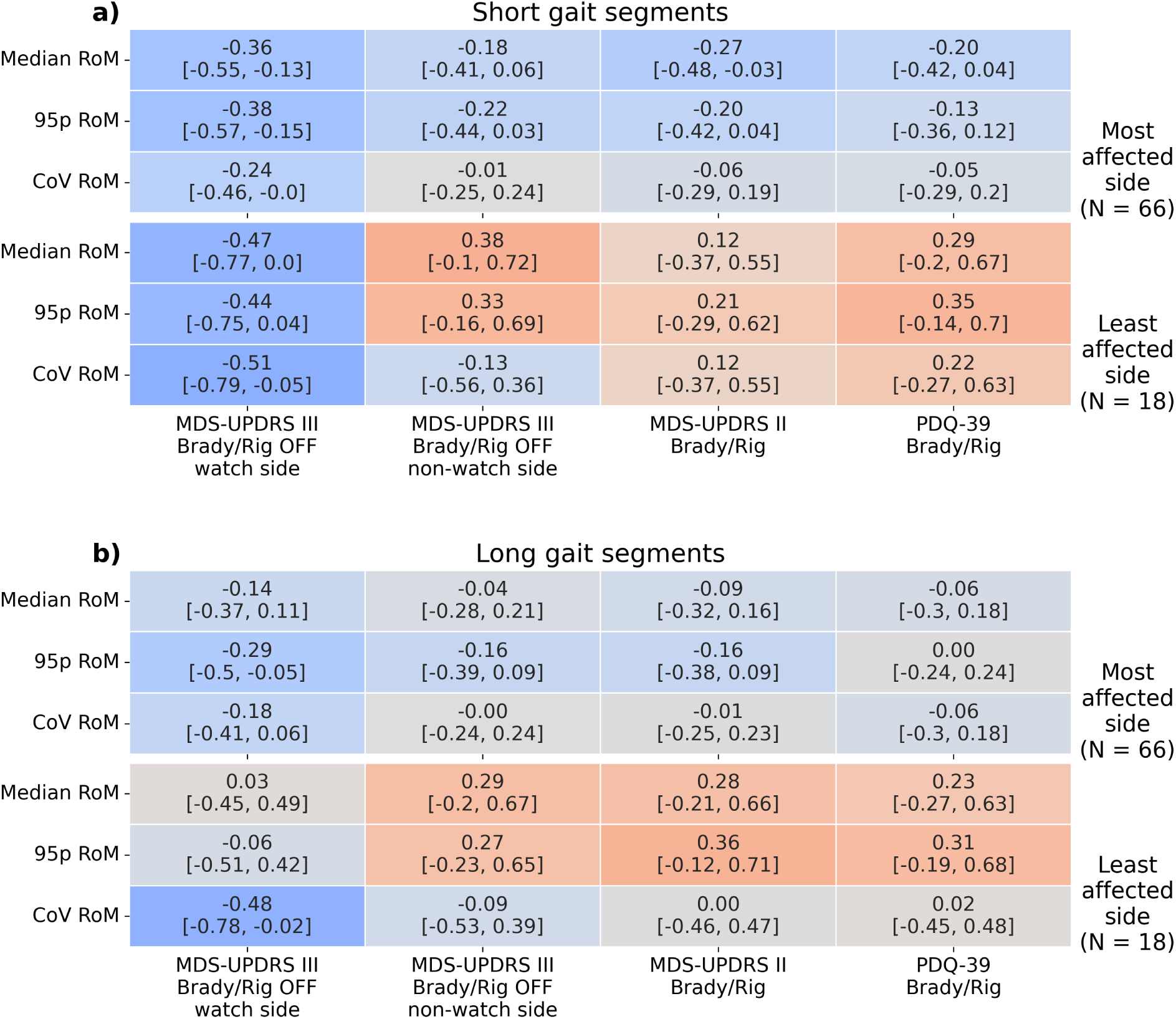
Associations between digital and clinical measures in unmedicated participants. Values represent Spearman rank correlations between the digital measures and clinical scores, with squared brackets representing 95% bias-corrected and accelerated bootstrap confidence intervals. Correlations are shown separately for **(a)** short gait segments (*<* 20 seconds) and **(b)** long gait segments (*<* 20 seconds). RoM: range of motion; 95p: 95^th^ percentile; CoV: coefficient of variation; MDS-UPDRS: Movement Disorder Society-sponsored Unified Parkinson’s Disease Rating Scale; PDQ-39: Parkinson’s Disease Questionnaire-39; MDS-UPDRS III Brady/Rig: the sum of MDS-UPDRS Part III unilateral bradykinesia and rigidity items (unilateral items of 3.3–3.8); MDS-UPDRS II Brady/Rig: the sum of MDS-UPDRS Part II bradykinesia and rigidity items (2.4–2.7); PDQ-39 Brady/Rig: the sum of PDQ-39 bradykinesia and rigidity items (11–16); watch side: unilateral items of the same side the watch is worn; non-watch side: unilateral items of the side opposite to where the watch is worn; OFF: after overnight withdrawal of dopaminergic medication; ON: one hour after taking dopaminergic medication.

Median and 95^th^ percentile range of motion were strongly correlated in both short (*r_s_* = 0.91) and long (*r_s_* = 0.85) gait segments (Supplementary Figure 6). CoV range of motion was moderately correlated with the other measures in short gait segments (*r_s_* = 0.58–0.62) and weakly correlated in long gait segments (*r_s_* = 0.26–0.41).

#### Test-retest reliability

Intraclass correlation coefficients (ICCs) between the second and third week of follow-up were high for both the most affected side (0.87–0.95) and least affected side (0.84–0.97), with generally slightly higher ICCs in short gait segments (Table 2).

**Table 2:** Test-retest reliability of digital measures. Values represent intraclass correlation coefficients (ICCs) with 95% bias-corrected and accelerated bootstrap confidence intervals for each arm swing measure and gait segment duration. ICCs are calculated between the second and third week of follow-up. Short: *<* 20 seconds; long: ≥ 20 seconds; perc: percentile; CoV: coefficient of variation; RoM: range of motion.

|  |  | Gait segment duration |  |
| --- | --- | --- | --- |
|  |  | Short | Long |
| PD most affected side | Median RoM | 0.95 [0.94, 0.96] | 0.91 [0.89, 0.93] |
|  | 95<sup>th</sup> perc RoM | 0.93 [0.91, 0.95] | 0.94 [0.92, 0.95] |
|  | Median CoV RoM | 0.94 [0.92, 0.95] | 0.87 [0.84, 0.90] |
| PD least affected side | Median RoM | 0.93 [0.90, 0.95] | 0.89 [0.85, 0.92] |
|  | 95<sup>th</sup> perc RoM | 0.97 [0.96, 0.98] | 0.92 [0.89, 0.94] |
|  | Median CoV RoM | 0.93 [0.90, 0.95] | 0.84 [0.78, 0.88] |

### Longitudinal

The two-year longitudinal analyses included 22 out of 98 unmedicated PD participants (17 most affected side, 5 least affected side), 301 out of 467 medicated PD participants (187 most affected side, 114 least affected side), and 29 out of 45 controls (Supplementary Figures 7 and 8). The baseline demographic and clinical characteristics are summarized in Table 3. As expected, the PD medicated group had higher MDS-UPDRS Part I and Part II scores compared with the unmedicated group (most affected side: *p* = 0.001; least affected side: *p* = 0.061). In two years, the amount of filtered gait in short gait segments decreased in unmedicated PD participants (*p* = 0.048), but not significantly in medicated PD participants (*p* = 0.169) and controls (*p* = 0.190; Figure 4). In long gait segments, the amount of filtered gait did not change in any group (unmedicated PD: *p* = 0.305; medicated PD: *p* = 0.316; controls: *p* = 0.917). For longitudinal changes in unfiltered gait quantity, see Supplementary Figure 9.

**Figure 4:**
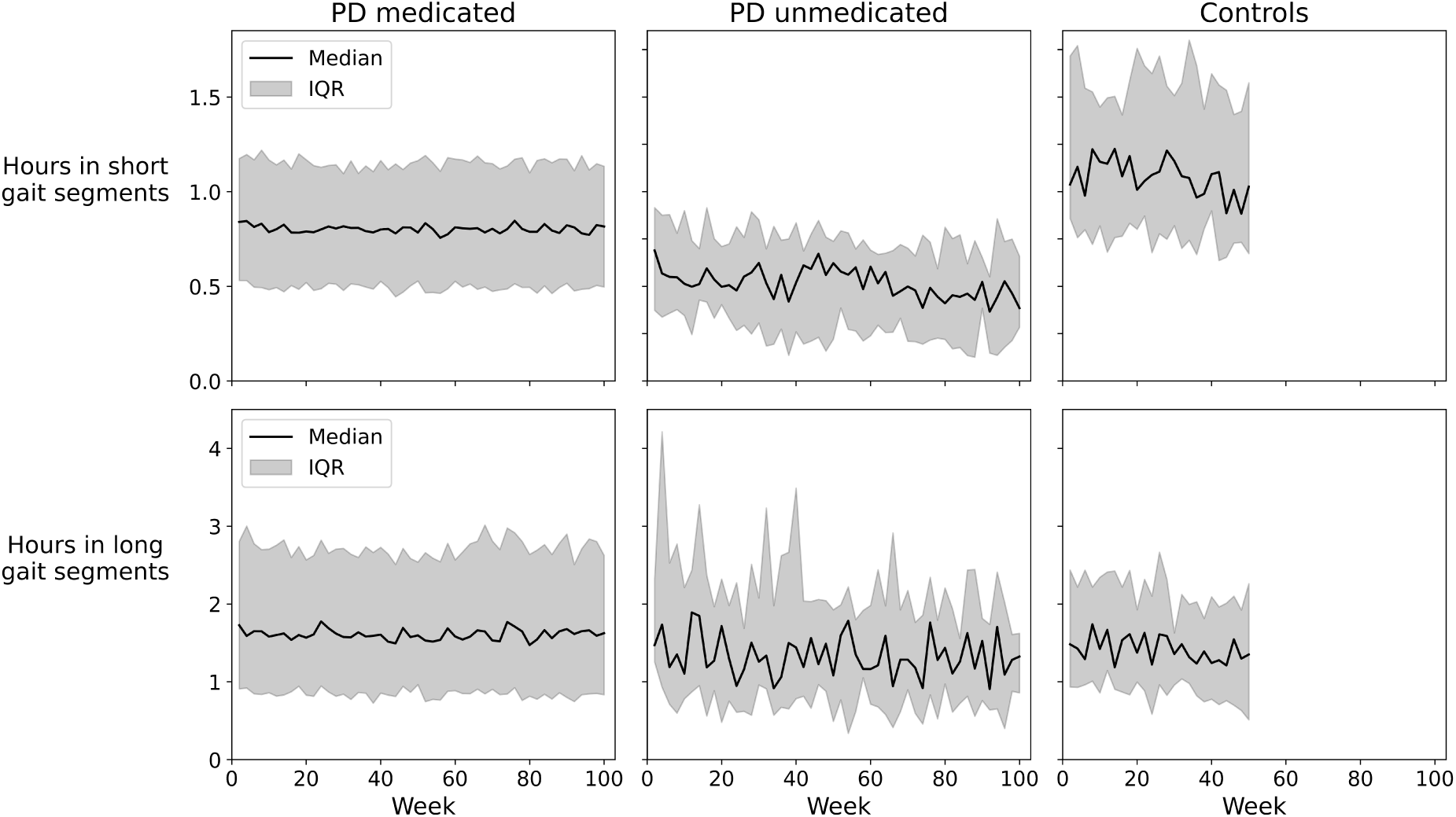
Weekly amount of gait without other arm activities. Quantities are shown for short (*<* 20 seconds) and long (≥ 20 seconds) gait segments aggregated across participants included in the two-year standardized response mean (SRM) analysis (i.e., those remaining in the study after two years). IQR: interquartile range. Using the *ℓ*_1_ trend filter, individual trends were estimated from the biweekly measurements (Figure 5). Longitudinal changes in the digital measures are presented in Supplementary Figures 10 and 11. Changes in median and 95^th^ percentile RoM were strongly correlated, whereas changes in CoV RoM showed moderate or no correlation with the other digital measures (Supplementary Figure 12).

**Figure 5:**
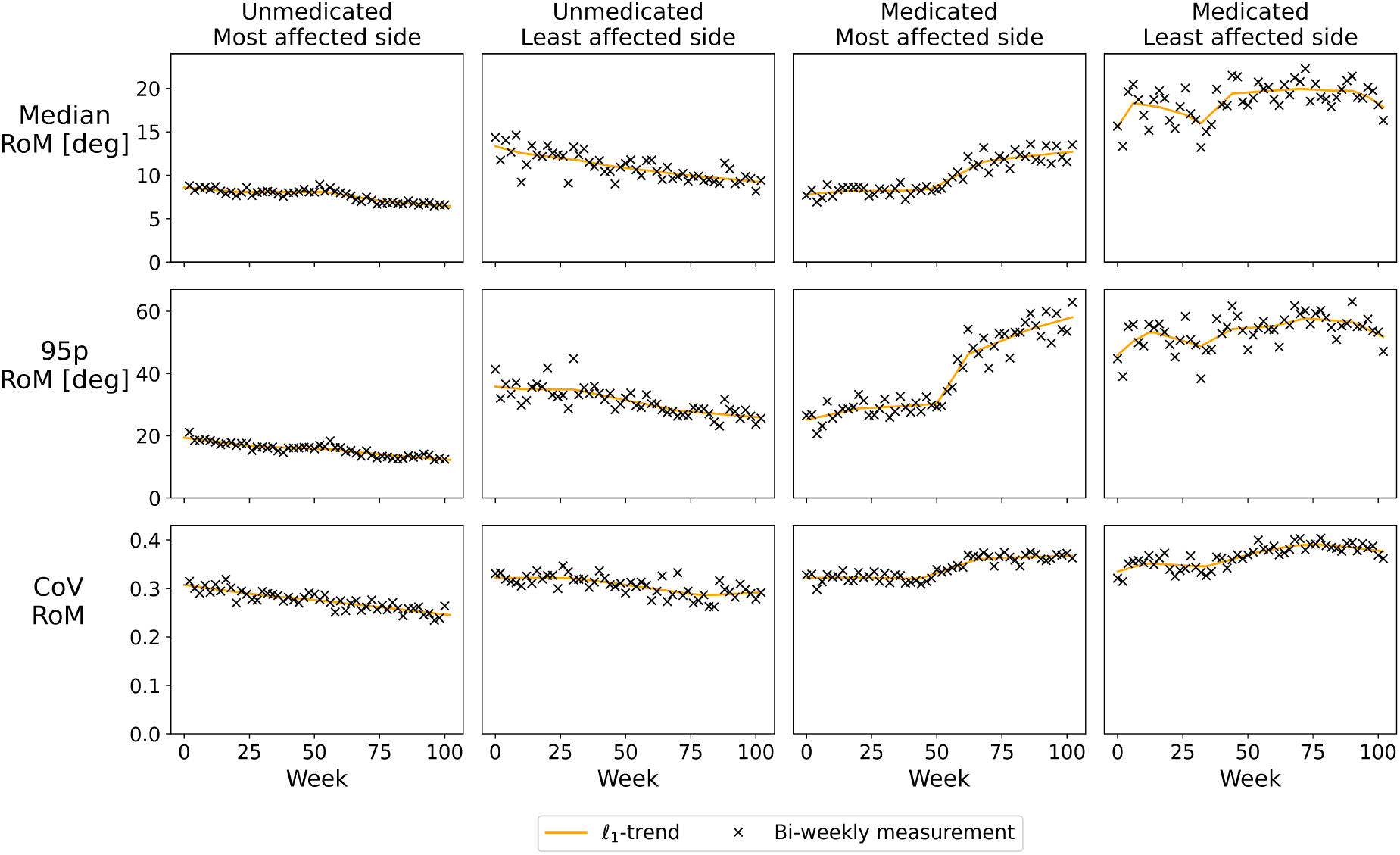
Example fit if *ℓ*_1_ trend filter. Each subfigure shows an illustrative examples of the *ℓ*_1_ trend filter applied to biweekly range of motion (RoM) measurements. Each column corresponds to a single participant, selected as representative of the overall group. The orange line shows the estimated trend and the black crosses indicate individual biweekly RoM values. 95p: 95^th^ percentile; CoV: coefficient of variation; deg: degrees.

**Table 3:** Demographics of the longitudinal study population. MAS: participants wearing the watch on the most affected side; LAS: participants wearing the watch on the least affected side; N: number of participants; SD: standard deviation; N/A: not applicable; MDS-UPDRS: Movement Disorder Society-sponsored Unified Parkinson’s Disease Rating Scale; Part III subscore: the sum of MDS-UPDRS Part III unilateral bradykinesia and rigidity items (unilateral items of 3.3—3.8).

|  | PD medicated |  | PD unmedicated |  | Controls |
| --- | --- | --- | --- | --- | --- |
|  | MAS<br>(N = 189) | LAS<br>(N = 118) | MAS<br>(N = 17) | LAS<br>(N = 5) | (N = 29) |
| Women (N (%)) | 69 (36.5) | 55 (46.6) | 9 (52.9) | 1 (20.0) | 16 (55.2) |
| Age (years, mean ± SD) | 61.8 ± 8.6 | 62.0 ± 7.6 | 61.9 ± 10.8 | 62.2 ± 16.0 | 70.7 ± 8.3 |
| Watch on dominant side (N (%)) | 38 (20.1) | 16 (13.6) | 5 (29.4) | 1 (20.0) | 7 (24.1) |
| **Hoehn & Yahr stage (N)** |  |  |  |  |  |
| I | 16 (8.5) | 8 (6.8) | 2 (11.8) | 1 (20.0) | N/A |
| II | 153 (81.0) | 98 (83.1) | 12 (70.6) | 4 (80.0) | N/A |
| III | 20 (10.6) | 12 (10.2) | 3 (17.6) | 0 (0.0) | N/A |
| **MDS-UPDRS (mean ± SD)** |  |  |  |  |  |
| Part I | 7.3 ± 3.4 | 7.0 ± 3.9 | 4.4 ± 3.6 | 3.6 ± 2.9 | N/A |
| Part II | 6.3 ± 4.7 | 5.4 ± 4.4 | 2.8 ± 2.5 | 2.6 ± 1.7 | N/A |
| Part III OFF | 23.6 ± 9.1 | 22.0 ± 8.8 | 22.1 ± 7.6 | 27.4 ± 4.6 | N/A |
| Part III ON | 20.9 ± 9.2 | 19.0 ± 8.6 | N/A | N/A | N/A |
| Part III subscore OFF watch side | 11.4 ± 4.0 | 7.2 ± 3.8 | 10.5 ± 3.1 | 8.8 ± 2.2 | N/A |
| Part III subscore ON watch side | 10.1 ± 4.1 | 6.6 ± 3.9 | N/A | N/A | N/A |

#### Sensitivity to change

Figure 6 shows the change in standardized response means (SRMs) of all digital measures across the two-year follow-up period, whereas Figure 7 shows the SRMs of all digital measures and the MDS-UPDRS part III subscores at year one and two.

**Figure 6:**
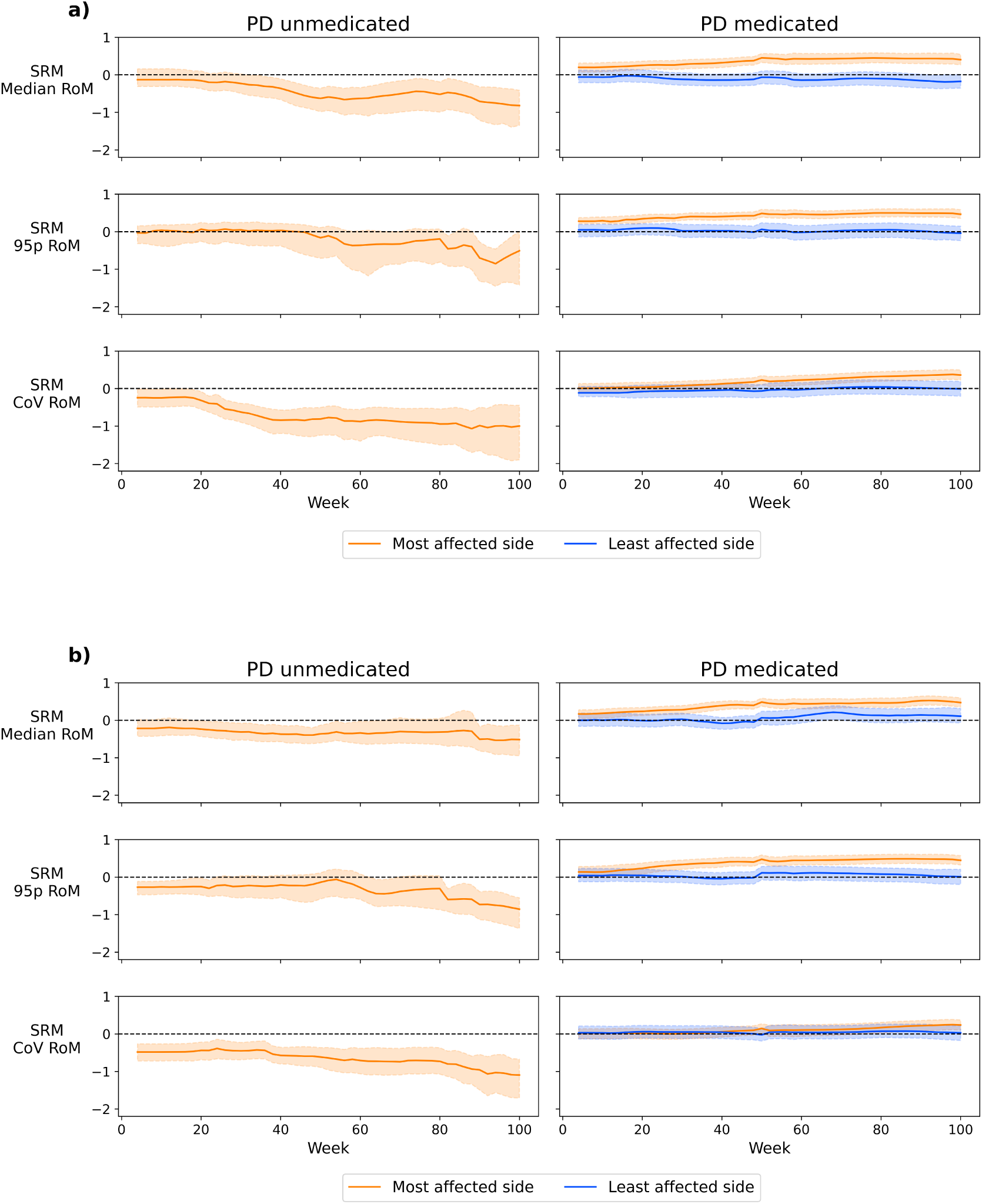
Sensitivity to change for varying study durations. Bold lines represent the standardized response means (SRMs) of the digital measures in **(a)** short gait segments (*<* 20 seconds) and **(b)** long gait segments (≥ 20 seconds). Shaded areas represent 95% bias-corrected and accelerated bootstrap confidence intervals. RoM: range of motion; 95p: 95^th^ percentile; CoV: coefficient of variation.

**Figure 7:**
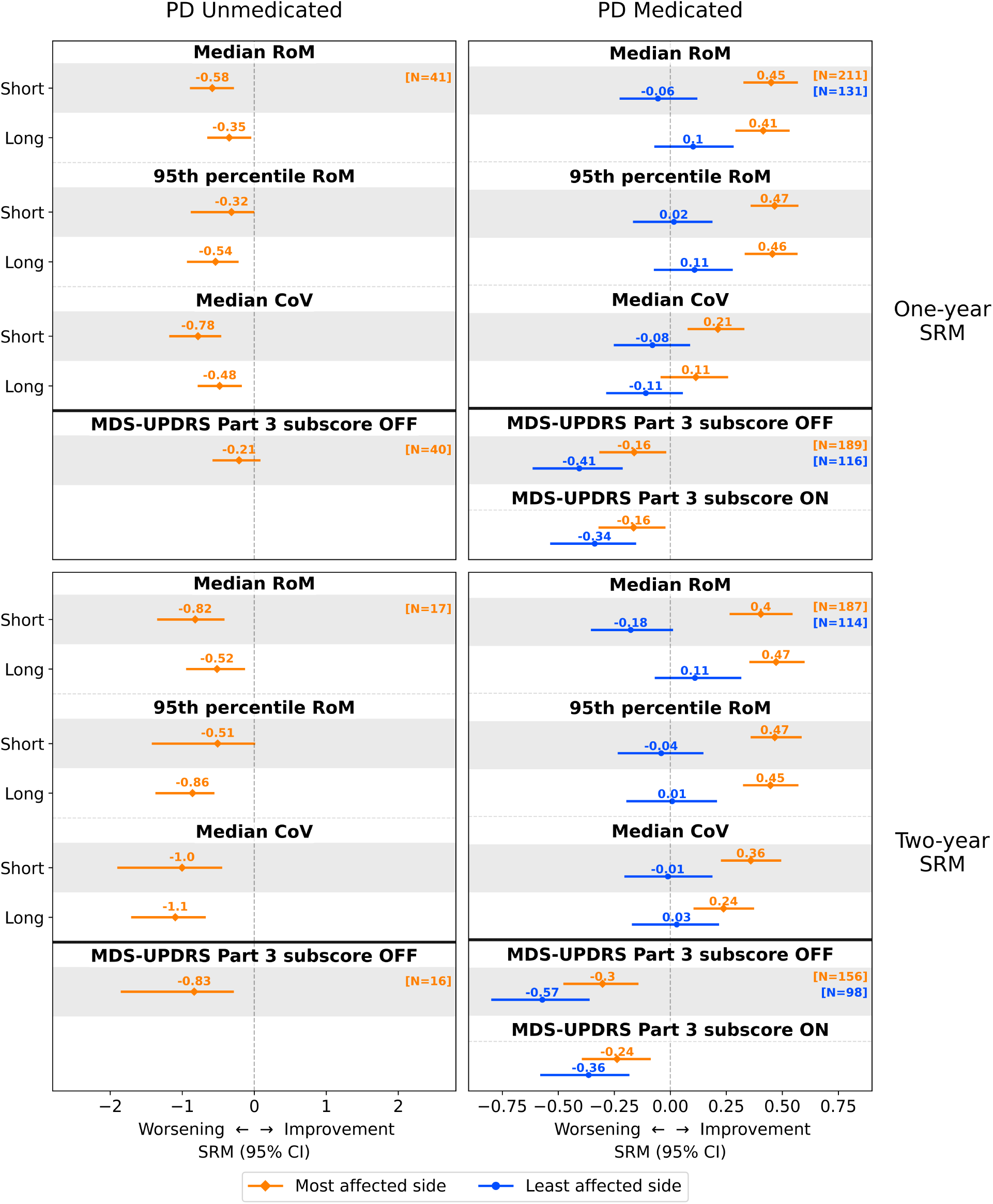
One- and two-year sensitivity to change of the digital measures and clinical scores. Values represent standardized response means (SRMs), error bars represent 95% bias-corrected and accelerated bootstrap confidence intervals (CIs). For comparability across measures, the MDS-UPDRS Part 3 scores were sign-flipped so that positive SRM values correspond to improvement and negative values to worsening. RoM: range of motion; CoV: coefficient of variation; MDS-UPDRS: Movement Disorder Society-sponsored Unified Parkinson’s Disease Rating Scale; Short: gait segments *<* 20 seconds; Long: gait segments ≥ 20 seconds.

In PD unmedicated, only the SRM of the most affected side was reported because of unreliable SRM estimates on the least affected side due to small sample size (N=5). On the most affected side, all digital measures decreased over one year (SRMs of -0.32 to -0.78), and all over two years (SRMs of -0.51 to -1.10). In contrast, the unilateral MDS-UPDRS Part III subscore OFF only worsened significantly over two years (one year SRM: -0.21; two year SRM: -0.83).

In PD medicated, all digital measures of the most affected side significantly increased over one year (SRMs of 0.11 to 0.47) and two years (SRMs of 0.24 to 0.47), apart from the CoV RoM in long gait segments at one year. Least affected side measures did not significantly change. By contrast, unilateral MDS-UPDRS Part III subscores in OFF and ON increased over one and two years for both the most and least affected sides. In controls, the median and 95^th^ percentile RoM remained stable over one year, whereas the CoV RoM decreased slightly (Supplementary Figure 13).

To confirm the robustness of our findings, we performed multiple sensitivity analyses. Not applying the *ℓ*_1_ trend filter to biweekly measures resulted in comparable direction and magnitude of changes with increased week-to-week variability (Supplementary Figure 14). Using unfiltered gait instead of filtered gait did not alter the conclusions (Supplementary Figures 15 and 16). Not applying IPCW produced comparable one-year SRMs, but resulted in an underestimation of two-year SRMs for most digital measures (Supplementary Figures 17 and 18).

#### Impact of dopaminergic medication

The observed two-year increase in digital measures on the most affected side in the PD medicated group was independently associated with both (1) an increase in LEDD, and (2) the development of slight dyskinesia (MDS-UPDRS Part IV item 4.1 = 1; Figure 8).

**Figure 8:**
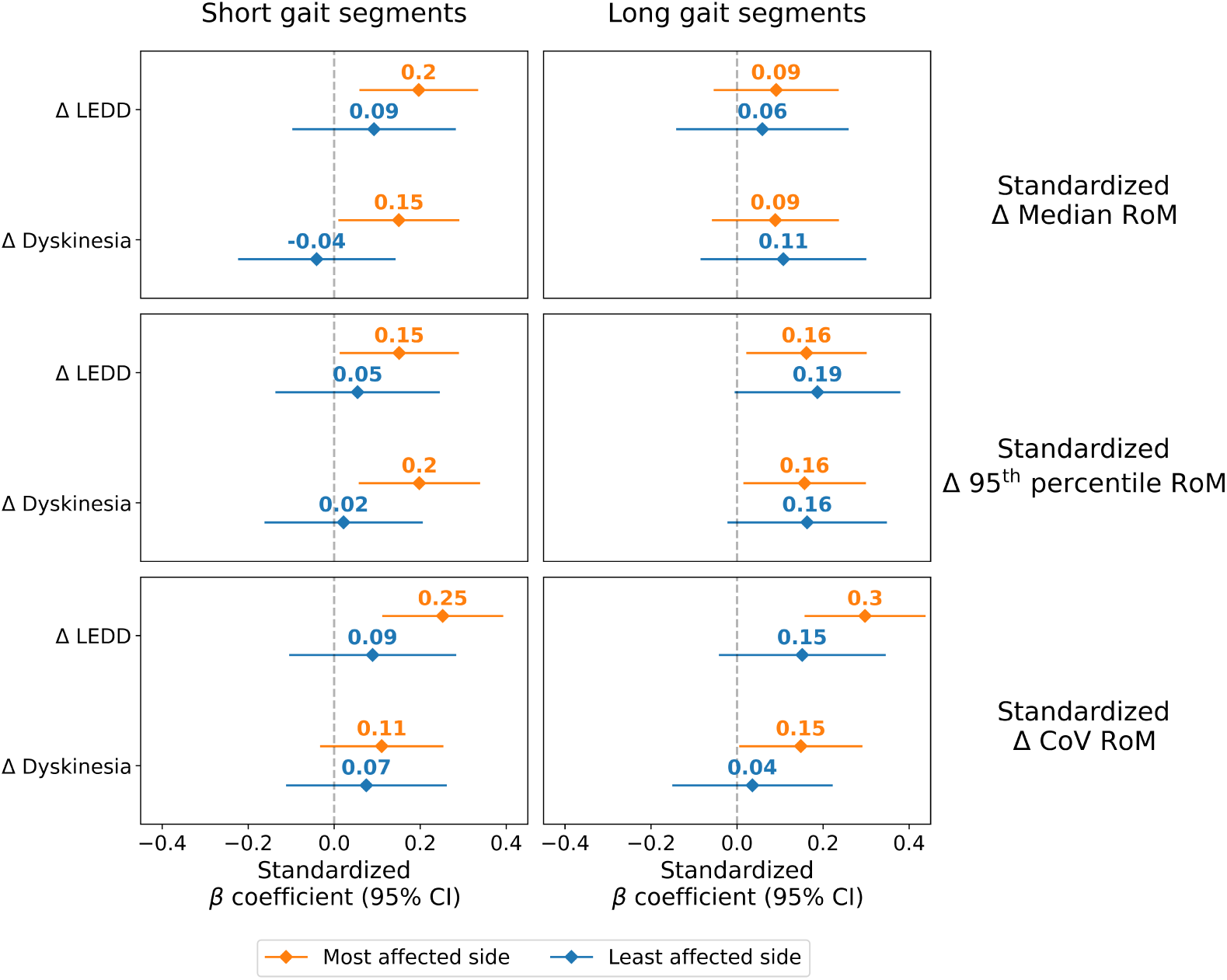
Effect of changes in medication on changes in digital measures. Values represent standardized coefficients of ordinary least squares (OLS) regression models with standardized two-year change in digital measures as dependent variable and disease characteristics and medication effects as covariates. The covariates are whether the watch is worn on the most affected side (watch side), two-year change in LEDD, interaction between the watch side and change in LEDD, change in dyskinesia, interaction between watch side and change in dyskinesia, months since diagnosis, and interaction between watch side and months since diagnosis. Coefficients represent a standard deviation change in the digital measure per standard deviation increase in the covariate. Coefficients are shown separately for most affected side (orange) and least affected side (blue). Error bars represent 95% bias-corrected and accelerated bootstrap confidence intervals (CIs). RoM: range of motion; CoV: coefficient of variation; LEDD: levodopa equivalent daily dose.

## Discussion

This study presents a longitudinal evaluation of digital arm swing measures obtained during free-living gait in early-stage PD, assessing construct validity, test-retest reliability, and sensitivity to change. The digital measures were reduced in PD compared to controls, were lower on the most affected side relative to the least affected side, and correlated moderately clinical evaluations of bradykinesia and rigidity. In addition, the digital measures demonstrated excellent week-to-week reliability, and were sensitive to disease progression in unmedicated participants, with effect sizes larger than those of clinical evaluations of bradykinesia and rigidity at one-year follow-up, and comparable to those at two-year follow-up. This one-year change of digital measures in unmedicated participants was larger than in controls, suggesting that observed changes were predominantly PD-related. In medicated PD, longitudinal improvements in arm swing were observed on the most affected side, which was associated with increases in LEDD and the development of upper-limb dyskinesia.

In unmedicated participants, arm swing declined over time, whereas medicated participants showed improvement on the most affected side and no changes on the least affected side. This pattern is likely attributable to dopaminergic therapy, which is known to improve arm swing during gait [22, 46, 47]. Prior studies have demonstrated that dopaminergic treatment can mask worsening of arm swing, even when measured in the OFF state [48]. It is therefore not unexpected that arm swing, measured under real-world conditions (which encompasses both ON and OFF states), would improve in the medicated group. Notably, unilateral MDS-UPDRS Part III subscores worsened in both ON and OFF states, suggesting that dopaminergic treatment may exert a relatively greater effect on arm swing compared to other fine motor symptoms, or, alternatively, that in-clinic assessments may not fully reflect at-home functioning (e.g., due to stress or fatigue). These findings suggest that digital arm swing measures are particularly informative endpoints for studying disease progression in early-stage, untreated PD. The clear presence of arm swing alterations at baseline in de novo patients further indicates their potential utility in prodromal cohorts, as arm swing reductions often precede a clinical diagnosis and may emerge before lower-limb impairments [49, 50]. After initiation of dopaminergic treatment, however, passive monitoring alone may be insufficient to capture disease progression, and may need to be complemented by in-clinic or active OFF-state assessments.

Few longitudinal studies have evaluated the sensitivity to change of arm swing measures in PD. A recent review of digital outcomes in early PD [13] identified only one study assessing longitudinal changes in arm swing, reporting a one-year decline in RoM during a 2-minute in-clinic walk task [48]. However, its small sample size (n = 7), controlled setting, and absence of standardization of outcomes limit generalizability. In a study postdating the review, a stronger one-year reduction in arm swing RoM was observed in the clinic compared to at home during a 1-minute walk task, suggesting that environmental variability may reduce sensitivity [24]. By contrast, our passively collected measures captured significant one-year declines in median, 95^th^ percentile, and CoV RoM in unmedicated participants across both short and long gait segments. The one-year standardized response means (SRMs) of these digital measures were comparable to those previously reported in both clinic- and home-based standardized arm swing measurements. Notably, the SRMs of single arm swing measures were larger than a composite clinical score based on clinical evaluation of bradykinesia and rigidity, despite composite scores often being more sensitive to change than individual items [24, 51, 52]. Together, these findings suggest that arm swing measures obtained through passive, real-world monitoring can capture longitudinal change with at least comparable sensitivity to MDS-UPDRS composite scores, supporting their potential as progression markers in early PD.

In addition to the median and 95^th^ percentile RoM, we examined the CoV RoM as a marker of arm swing variability. Whereas previous studies of lower-limb gait variability in free-living conditions have reported mixed findings, most observed increased variability in individuals with PD [53]. In contrast, we found reduced arm swing variability in PD compared to controls, lower variability on the most affected side compared with the least affected side, and further reductions over time in unmedicated PD. This discrepancy may reflect genuine differences between measuring gait at the upper and lower limbs, as our results align with earlier observations of increased lower-limb but reduced upper-limb variability in free-living gait in PD [54]. This may be partly explained by reduced passive movement in the upper limbs due to rigidity. Additionally, lower variability in the upper limbs may reflect reduced postural adjustments in response to the environment, leading to more stereotyped movement patterns. Impaired arm-leg synchronization during gait in PD may contribute to this discrepancy [55].

Whereas the median and 95^th^ percentile RoM were highly correlated, both measures showed only moderate correlations with arm swing variability, suggesting that this measure may partly capture distinct aspects of motor impairment. In the unmedicated cohort, all digital measures were inversely associated with bradykinesia and rigidity scores on the watch side. In contrast, in the medicated cohort, only the median and 95^th^ percentile RoM retained consistent associations, whereas variability was not significantly correlated with bradykinesia and rigidity evaluations. Moreover, variability improved less over time in the medicated cohort compared to the other measures. One possible explanation is that variability reflects aspects of postural control that are predominantly mediated by nondopaminergic pathways and therefore less responsive to dopaminergic therapy [47, 56–58]. In addition, behavioral factors–particularly gait speed–may also influence arm swing variability, as lower gait speed is associated with increased gait variability [59, 60]. This may contribute to the observed association between greater self-reported gait difficulties and increased variability, as patients who experience more gait difficulties or fear of falling may adapt by decreasing their gait speed. In conclusion, arm swing variability is a promising digital biomarker as it showed high sensitivity to progression in the unmedicated cohort, but more research is needed to understand its clinical interpretation when measured in free-living conditions, for example by including more elaborate balance assessments and gait speed measurements.

Whether short or long gait segments best capture PD-related changes has been debated [27, 61, 62]. Prior work showed that medication-induced increases in arm swing were most pronounced during longer gait segments [22], leading us to hypothesize that arm swing during longer segments would be more sensitive to change. Unexpectedly, however, both short and long segments demonstrated clear lateral differences and sensitivity to change. This highlights that short gait segments—though often overlooked—can meaningfully contribute to tracking disease progression in real-world settings. One possible explanation is that gait initiation and termination, which are disproportionally represented in shorter segments and are known to be impaired in PD, amplify the differences observed in these gait segments [50, 63].

In a previous cross-sectional study, filtering gait (i.e., discarding gait segments with other arm activities) increased the medication-induced change in both median and 95^th^ percentile RoM [22]. In the current longitudinal study, however, filtering did not substantially improve sensitivity to change. This is not entirely unexpected, as non-gait upper-limb movements also decline in PD, suggesting that reduced RoM may be a generalized motor feature rather than restricted to arm swing during gait [14]. Thus, filtering gait may primarily serve to enhance the specificity and interpretability of digital measures, rather than to improve their sensitivity to change.

Digital arm swing measures demonstrated excellent test-retest reliability, with ICCs ranging from 0.93 to 0.97 in gait segments under 20 seconds and from 0.87 to 0.94 in longer segments. In a prior study using a 2-minute 10-meter walk test, month-to-month in-clinic ICC was 0.84, and two-week at-home ICCs ranged from 0.77 to 0.92 [24]. In another study involving a weekly up-and-go test performed freely within the home environment, arm swing acceleration had a week-to-week ICC of just 0.43, suggesting that task complexity may reduce reliability [6]. Interestingly, the excellent reliability observed in our passively collected data suggests that weekly aggregated arm swing measures are relatively robust to short-term behavioral or environmental variations and can consistently capture arm swing in free-living conditions. Reliability was also higher than the reliability of the MDS-UPDRS Part III and its subscores for bradykinesia and rigidity (two-week ICCs from ranging from 0.85 to 0.89) [64, 65].

A final intriguing finding is that in participants wearing the watch on the least affected side, a larger median and 95^th^ percentile range of motion was associated with a higher sum of unilateral MDS-UPDRS Part III bradykinesia and rigidity items on the contralateral (most affected) side. This is essentially the inverse of the association observed on the watch side, and we suspect this is evidence of a compensatory mechanism: individuals with greater motor impairment on the most affected side may compensate by increasing arm swing on the least affected side. This interpretation aligns with prior reports of functional compensation by the least affected side for balance control in the lower limbs [66, 67]. In other words, enhanced movement on the least affected side may reflect a compensatory process aimed at stabilizing motor performance despite asymmetric motor impairment. Such compensation may be unconscious or, alternatively, an active strategy [68]

Several limitations warrant consideration. First, the two-year unmedicated group was small, as most participants initiated dopaminergic therapy during follow-up. While we observed consistent changes within this group, replication in larger cohorts where treatment initiation is postponed or in prodromal cohorts is needed. Second, controls were on average older than participants with PD. Although age was adjusted for in all cross-sectional analyses, residual age effects may have influenced longitudinal comparisons. Third, despite the use of reweighting to mitigate censoring bias, unmeasured factors may still have introduced residual bias. Nevertheless, this approach presents an improvement over prior work that did not account for censoring bias when studying PD progression. This study also has several notable strengths. The large sample size and exceptionally high long-term compliance with using a wearable device over two years provided a unique opportunity to objectively quantify long-term, within-subject changes in arm swing. We applied a validated, open-source algorithm for detecting and quantifying arm swing during free-living gait, promoting transparency and reproducibility in contrast to studies using proprietary methods. Moreover, the continuous, passive data collection across multiple years provides an ecologically valid view of motor function, capturing subtle, real-world disease progression that may be missed by episodic in-clinic testing.

## Author Contributions

EP, NT, LE and TL developed the methodology. EP processed and analyzed the data. LE, TH and BB supervised the study. YR, ML, JN, TH and KCH provided domain expertise. EP wrote the first version of the manuscript with LE and TL. All authors contributed to the editing of the paper and approved the final version of the manuscript.

## Acknowledgements

We are grateful to all participants and assessors of the Personalized Parkinson Project; without their participation and contributions, this study would not have been possible.

## Data Availability

Data from the Personalized Parkinson Project used in the present study were retrieved from the PEP database (https://pep.cs.ru.nl/index.html). The PPP data is available upon request via. More details on the procedure can be found on the website https://personalizedparkinsonproject.com/home. The code used for processing and analyzing the data is available at https://github.com/AI-for-Parkinson-Lab/longitudinal_arm_swing.

