## Supplementary material for "Longitudinal progression of digital arm swing measures during free-living gait in early Parkinson’s disease"

### Supplementary materials

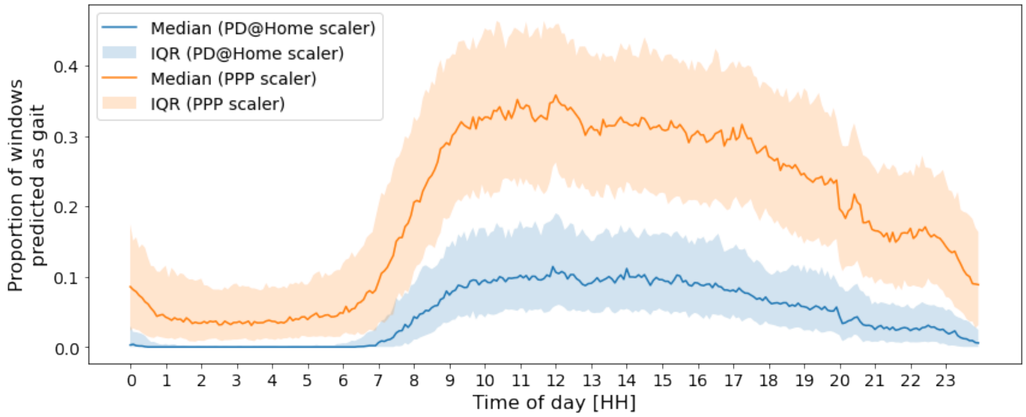


**Supplementary Figure 1: Daily gait distribution across participants.** To select an appropriate scaling method, we applied two different strategies to scale the classifier features: using a scaler fitted on the dataset on which the gait detection classifier was trained (PD@Home scaler) and a scaler fitted on participants of the Personalized Parkinson Project (PPP) dataset that adhere to the inclusion criteria of the original study (PPP scaler). Fitting a new scaler (PPP scaler) resulted in an unrealistically larger proportion of gait, likely attributable to a higher false positive rate. Using the PD@Home scaler resulted in a more expected daily proportion of gait [1, 2]. IQR: interquartile range.

**Supplementary Table 1: Generalizability of the gait detection pipeline.** To numerically evaluate the generalizability of the gait detection pipeline, we applied a manual annotation procedure. In this approach, one of the researchers (E. Post) "trained" to visually classify spectrograms of 2.000 windowed time series as either gait or non-gait using the wrist sensor data and video annotations of the Parkinson@Home Validation Study (PD@Home) as a training set. After training, we randomly sampled 1.000 predicted gait and 1.000 predicted non-gait windows from the Personalized Parkinson Project (PPP) dataset, with predictions based on the gait detection classifier. The "trained" researcher manually annotated these windows as gait or non-gait. Finally, the predictions of the classifier and the annotations of the researcher (serving as ground-truth) were compared to determine the classifier's performance (uncorrected). The evaluation metrics were corrected for the predicted prevalence of gait in the PPP dataset. We selected the PD@Home scaler due to its similarity in performance to the PD@Home dataset, in combination with demonstrating expected daily gait distributions in 1. Values are presented as mean (SD). We selected the PD@Home scaler due to its similarity in performance to the PD@Home dataset, in combination with demonstrating expected daily gait distributions in Supplementary Figure 1. PD@Home: a scaler fitted on the Parkinson@Home Validation study, the dataset on which the gait detection clasifier was trained; PPP subset: a scaler fitted on a subset of the PPP dataset clinically similar to the PD@Home study; PPP full: a scaler fitted on all participants of the PPP study; N/A: not applicable.

| | | | | Test set | | | |
|---|---|---|---|---|---|---|---|
| | | Validation set | | Uncorrected | | Corrected | |
| | | Sensitivity | Specificity | Sensitivity | Specificity | Sensitivity | Specificity |
| Scaler | PD@Home | 0.97 (0.05) | 0.88 (0.14) | 0.95 (0.10) | 0.74 (0.19) | 0.75 (0.24) | 0.94 (0.05) |
| | PPP subset | N/A | N/A | 0.92 (0.16) | 0.81 (0.13) | 0.68 (0.21) | 0.96 (0.02) |
| | PPP full | N/A | N/A | 0.89 (0.18) | 0.76 (0.17) | 0.58 (0.24) | 0.92 (0.03) |

**Supplementary Table 2: The list of MDS-UPDRS Part III categories included in the composed MDS-UPDRS Part III subscore.**

| Item | Name |
| --- | --- |
| Right Side | |
| 3.3b | Rigidity right upper extremity |
| 3.4a | Finger tapping right hand |
| 3.5a | Movement right hand |
| 3.6a | Pronation-supination right hand |
| 3.7a | Toe tapping right foot |
| 3.8a | Leg agility right leg |
| Left Side | |
| 3.3c | Rigidity left upper extremity |
| 3.3d | Rigidity right lower extremity |
| 3.4b | Finger tapping left hand |
| 3.5b | Movement left hand |
| 3.6b | Pronation-supination left hand |
| 3.7b | Toe tapping left foot |
| 3.8b | Leg agility left leg |

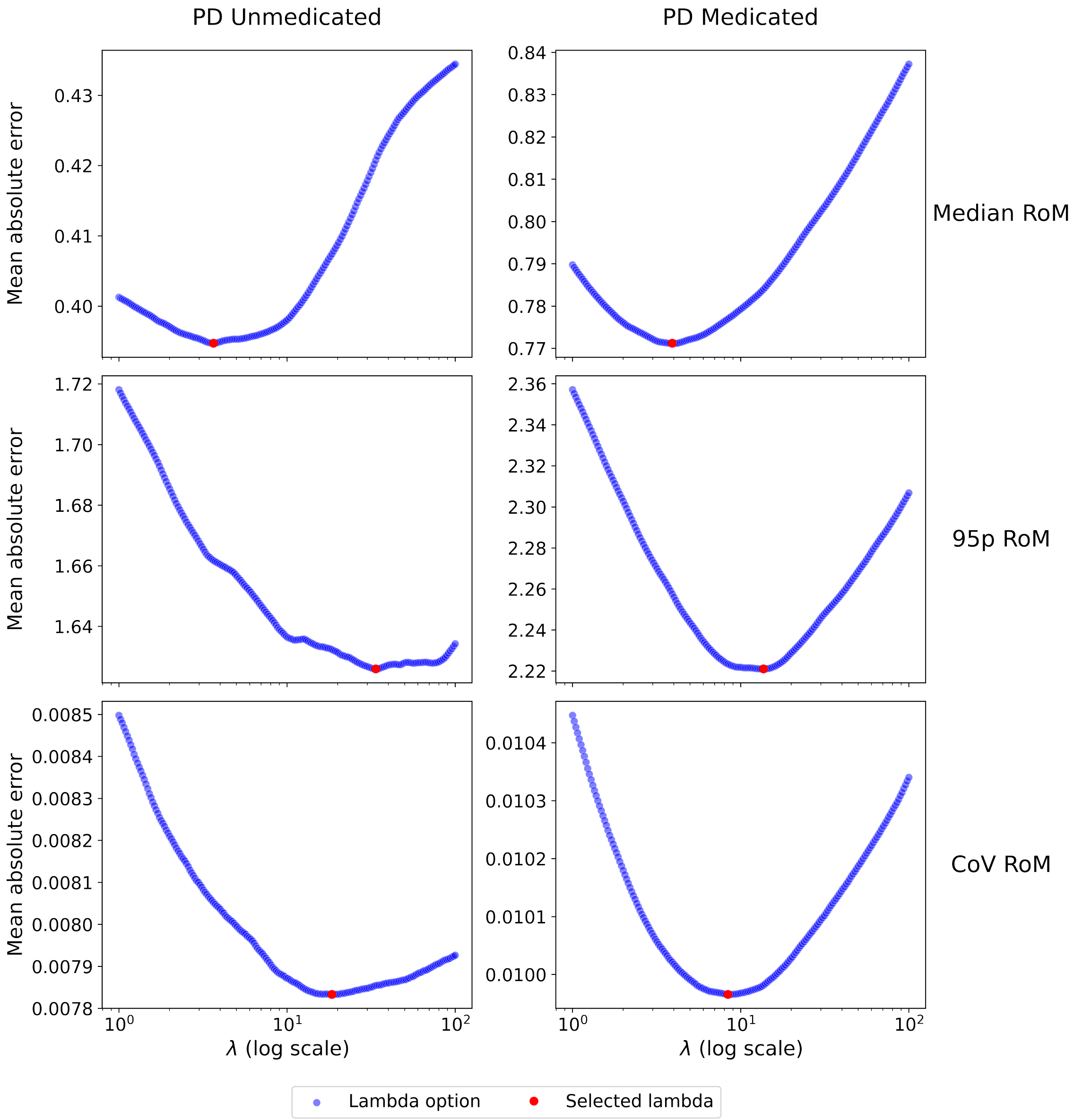


**Supplementary Figure 2: Validity of selected regularization parameters in $\ell_1$ trend filtering.** Each subfigure displays the mean absolute error of the $\ell_1$ trend filter for varying regularization parameter ($\lambda$) values of a digital measure. The red marker indicates the optimal $\lambda$ value, which was used for fitting an $\ell_1$ trend per participant applied in subsequent analyses. RoM: range of motion; 95p: $95^{\text{th}}$ percentile; CoV: coefficient of variation.

**Supplementary Table 3: Reliability of the extracted $\ell_1$ trends.** Values represent the median [interquartile range (IQR)] of the standard deviation (SD) of the signal, the SD of the residuals, and the signal-to-noise ratio (SNR) for the $\ell_1$ trend filter applied to the digital measures. The SNR is calculated as the ratio of the SD of the signal to the SD of the residuals. It represents the proportion of variance explained by the fitted trend. Note that coefficient of variation (CoV) range of motion (RoM) values are in % (divided by 100).

| | | Statistic | | |
|---|---|---|---|---|
| Parameter | Group | SD signal | SD residuals | SNR |
| Median RoM | PD Medicated | 2.2 [1.1, 3.8] | 3.2 [1.8, 4.7] | 0.7 [0.5, 1.0] |
| | PD unmedicated | 1.1 [0.6, 2.2] | 1.8 [1.2, 2.8] | 0.7 [0.4, 1.0] |
| | Controls | 1.1 [0.3, 2.6] | 3.1 [2.2, 5.1] | 0.4 [0.1, 0.5] |
| $95^{th}$ percentile RoM | PD Medicated | 3.5 [2.1, 5.3] | 5.0 [3.6, 6.7] | 0.7 [0.5, 1.0] |
| | PD unmedicated | 2.4 [1.3, 4.6] | 3.7 [2.5, 4.8] | 0.7 [0.4, 1.1] |
| | Controls | 2.9 [1.2, 3.5] | 4.6 [3.8, 5.6] | 0.5[ 0.3, 0.7] |
| CoV RoM (%) | PD Medicated | 0.8 [0.5, 1.3] | 1.3 [1.0, 1.5] | 0.6 [0.4, 1.0] |
| | PD unmedicated | 0.8 [0.5, 1.2] | 1.3 [1.1, 1.7] | 0.5 [0.3, 0.8] |
| | Controls | 0.3 [0.2, 0.5] | 0.9 [0.8, 1.3] | 0.3 [0.1, 0.6] |

| | PD | Controls |
|---|---|---|
| **Starting participants** | **621** | **50** |
| **Clinical exclusion criteria** | | |
| Walking aid | 58 | 0 |
| At least significant dyskinesia | 15 | N/A |
| **Remaining after clinical criteria** | **551 (89%)** | **50 (100%)** |
| **Measurement exclusion criteria** | | |
| Less than 3 days with 10 hours of sensor data between 08:00 and 22:00 | 66 | 4 |
| No filtered gait | 6 | 0 |
| No long gait segments | 7 | 1 |
| Less than 2 days with at least 2 minutes of filtered gait in first 3 and/or final 3 weeks | 45 | 1 |
| **Remaining after measurement criteria** | **427 (69%)** | **44 (88%)** |
| Watch on most affected side | 264 (62%) | N/A |
| Watch on least affected side | 156 (37%) | N/A |

**Supplementary Figure 3: Flowchart of cross-sectional participant selection.** Numbers in purple boxes denote participants included at each step, while numbers in white boxes denote participants excluded.

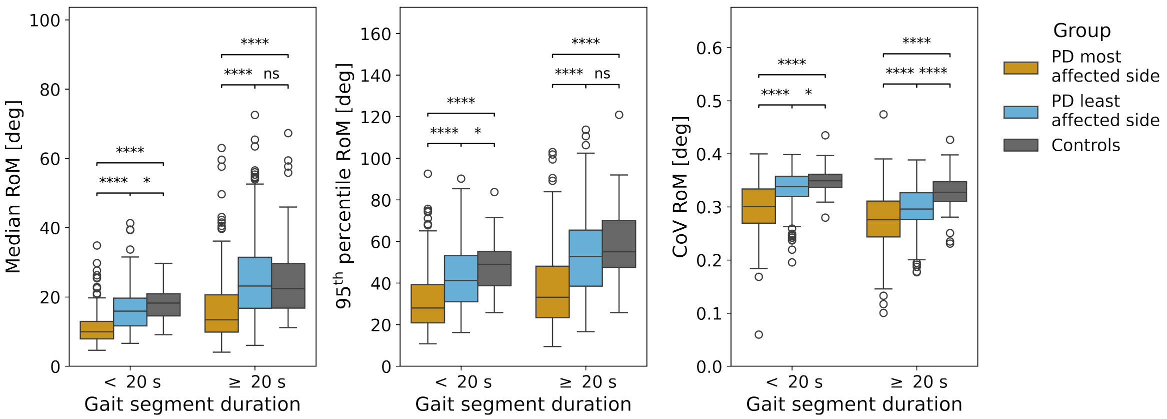


**Supplementary Figure 4: Unadjusted between-group differences of digital measures**. Comparisons were made using the non-parametric Wilcoxon rank-sum test of independent samples with 95% bias-corrected and accelerated bootstrap confidence intervals. Notation for two-sided thresholds of statistical significance: $p < 0.05$ (*), $p < 0.01$ (**), $p < 0.001$ (***), and $p < 0.0001$ (****): RoM: range of motion; CoV: coefficient of variation; deg: degrees; ns: not significant; short gait segments: $< 20$ seconds; long gait segments: $\geq 20$ seconds.

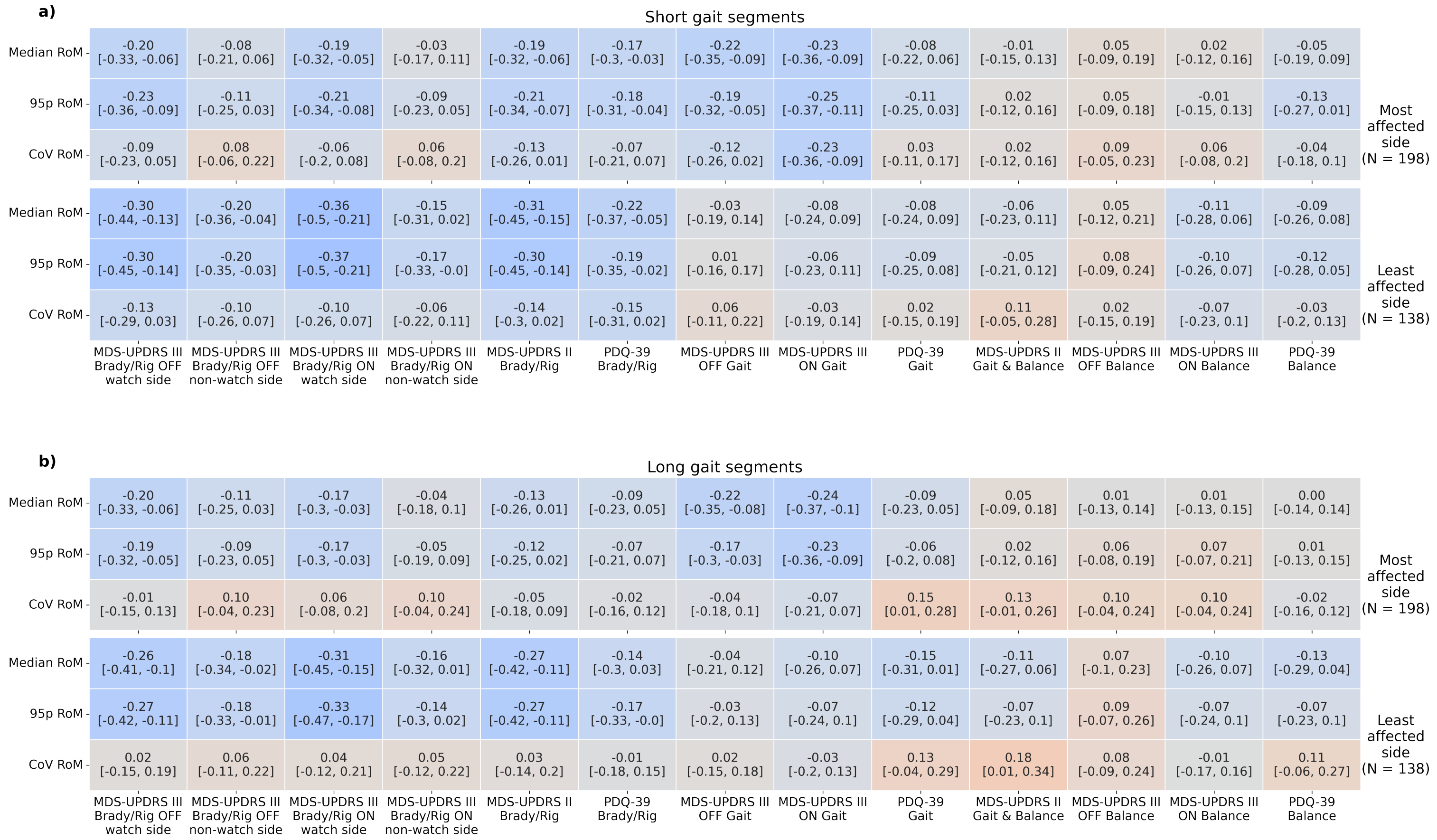


**Supplementary Figure 5: Associations between digital and clinical measures in medicated participants**. Values represent Spearman rank correlations between the digital measures and clinical scores (95% bias-corrected and accelerated bootstrap confidence intervals between squared brackets). Correlations are shown separately for **(a)** short gait segments ($<$ 20 seconds) and **(b)** long gait segments ($<$ 20 seconds). RoM: range of motion; 95p: 95$^{th}$ percentile; CoV: coefficient of variation; MDS-UPDRS: Movement Disorder Society-sponsored Unified Parkinson's Disease Rating Scale; PDQ39: Parkinson's Disease Questionnaire-39; UPDRS III Brady/Rig: the sum of MDS-UPDRS Part III unilateral bradykinesia and rigidity items(unilateral items of 3.3–3.8); UPDRS II Brady/Rig: the sum of MDS-UPDRS Part II bradykinesia and rigidity items (2.4–2.7); PDQ-39 Brady/Rig: the sum of PDQ-39 bradykinesia and rigidity items (11–16); UPDRS III Gait: MDS-UPDRS Part III item 3.10; PDQ-39 Gait: the sum of PDQ-39 gait items (4–6); UPDRS II Gait & Balance: MDS-UPDRS Part II item 2.12; UPDRS III Balance: MDS-UPDRS Part III item 3.12; PDQ-39 Balance: PDQ-39 item 9; watch side: unilateral items of the same side the watch is worn; non-watch side: unilateral items of the side opposite to where the watch is worn; OFF: after overnight withdrawal of dopaminergic medication; ON: one hour after taking dopaminergic medication.

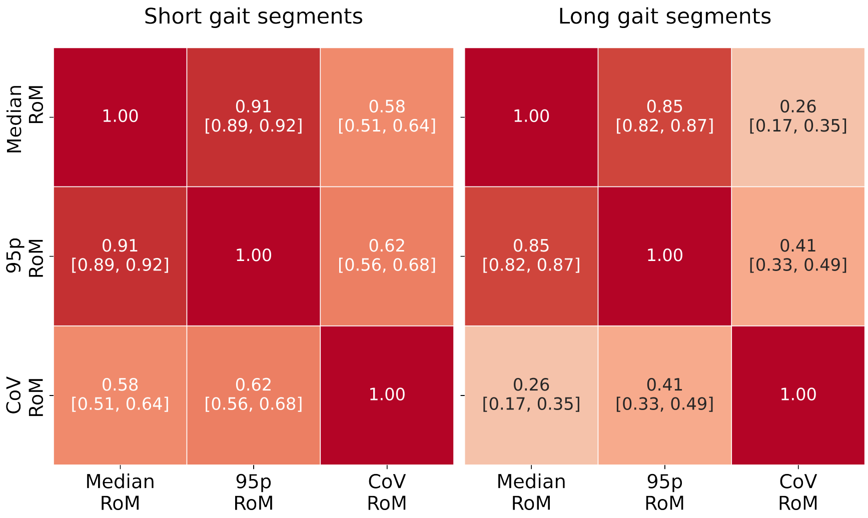


**Supplementary Figure 6: Association between digital measures.** Values represent Spearman rank correlations between the digital measures in short ($<$ 20 seconds) and long ($\geq$ 20 seconds) gait segments. Values between squared brackets represent 95% bias-corrected and accelerated bootstrap confidence intervals. RoM: Range of motion; 95p: 95$^{\text{th}}$ percentile; CoV: coefficient of variation.

| | PD medicated | PD unmedicated | Controls |
|---|---|---|---|
| **Starting participants** | **467** | **98** | **45** |
| **Clinical exclusion criteria** | | | |
| Starting medication | N/A | 59 | N/A |
| Walking aid | 98 | 0 | 0 |
| At least significant dyskinesia | 44 | N/A | N/A |
| Alternative diagnosis | 14 | 1 | N/A |
| Watch side switched | 14 | 11 | 1 |
| **Remaining after clinical criteria** | **332 (71%)** | **32 (33%)** | **44 (98%)** |
| **Measurement exclusion criteria** | | | |
| Less than 3 days with 10 hours of sensor data between 08:00 and 22:00 in first 3 and/or final 3 weeks | 16 | 6 | 16 |
| Less than 2 days with at least 2 minutes of filtered gait in first 3 and/or final 3 weeks | 7 | 1 | 0 |
| **Remaining after measurement criteria** | **309 (66%)** | **25 (26%)** | **29 (58%)** |
| Watch on most affected side | 189 (61%) | 18 (72%) | N/A |
| Watch on least affected side | 118 (38%) | 5 (20%) | N/A |

**Supplementary Figure 7: Flowchart of longitudinal participant selection.** Numbers in purple boxes denote participants included at each step, while numbers in white boxes denote participants excluded.

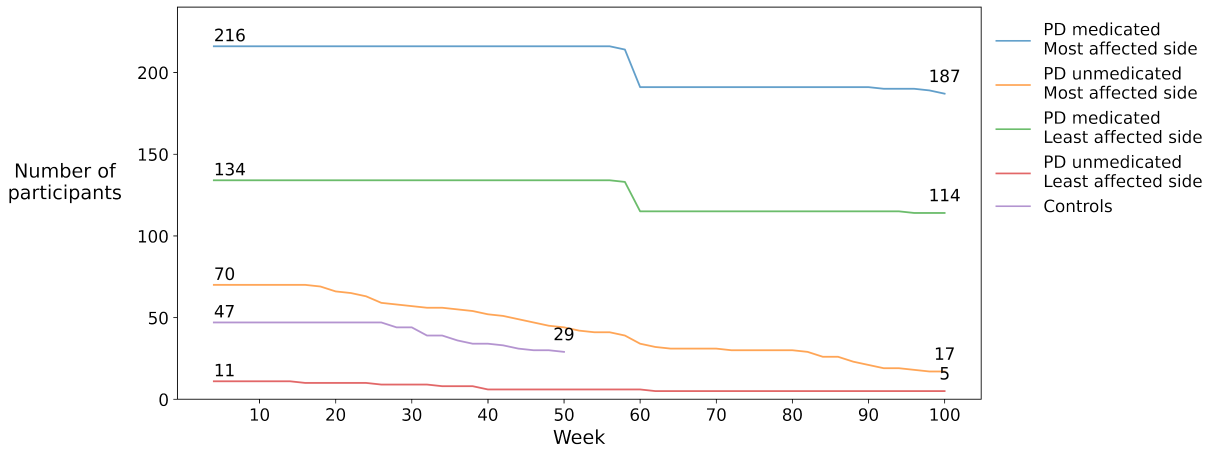


**Supplementary Figure 8: Survival curves per group.** Each line denotes the number of participants remaining in the study for varying study durations. Values above the curves denote group size, with the first number indicating participants at baseline and the second number indicating participants remaining after two years.

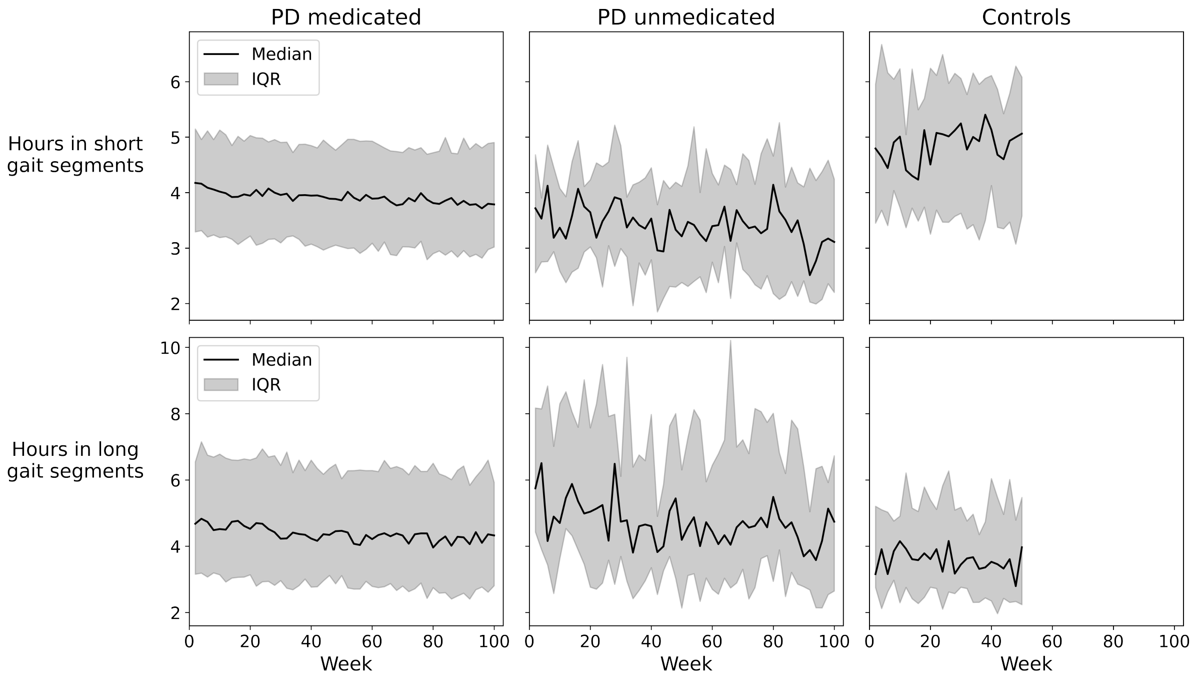


**Supplementary Figure 9: Weekly amount of unfiltered gait**. Quantities are shown for short (< 20 s) and long ($\geq$ 20 s) gait segments aggregated across participants included in the two-year SRM analysis (i.e., those remaining in the study after two years). The amount of gait decreased in medicated PD (short gait segments: $p < 0.001$; long gait segments: $p < 0.01$), but not in unmedicated PD (short gait segments: $p = 0.08$, long gait segments: $p = 0.50$) or controls (short gait segments: $p = 0.86$; long gait segments: $p = 0.58$).

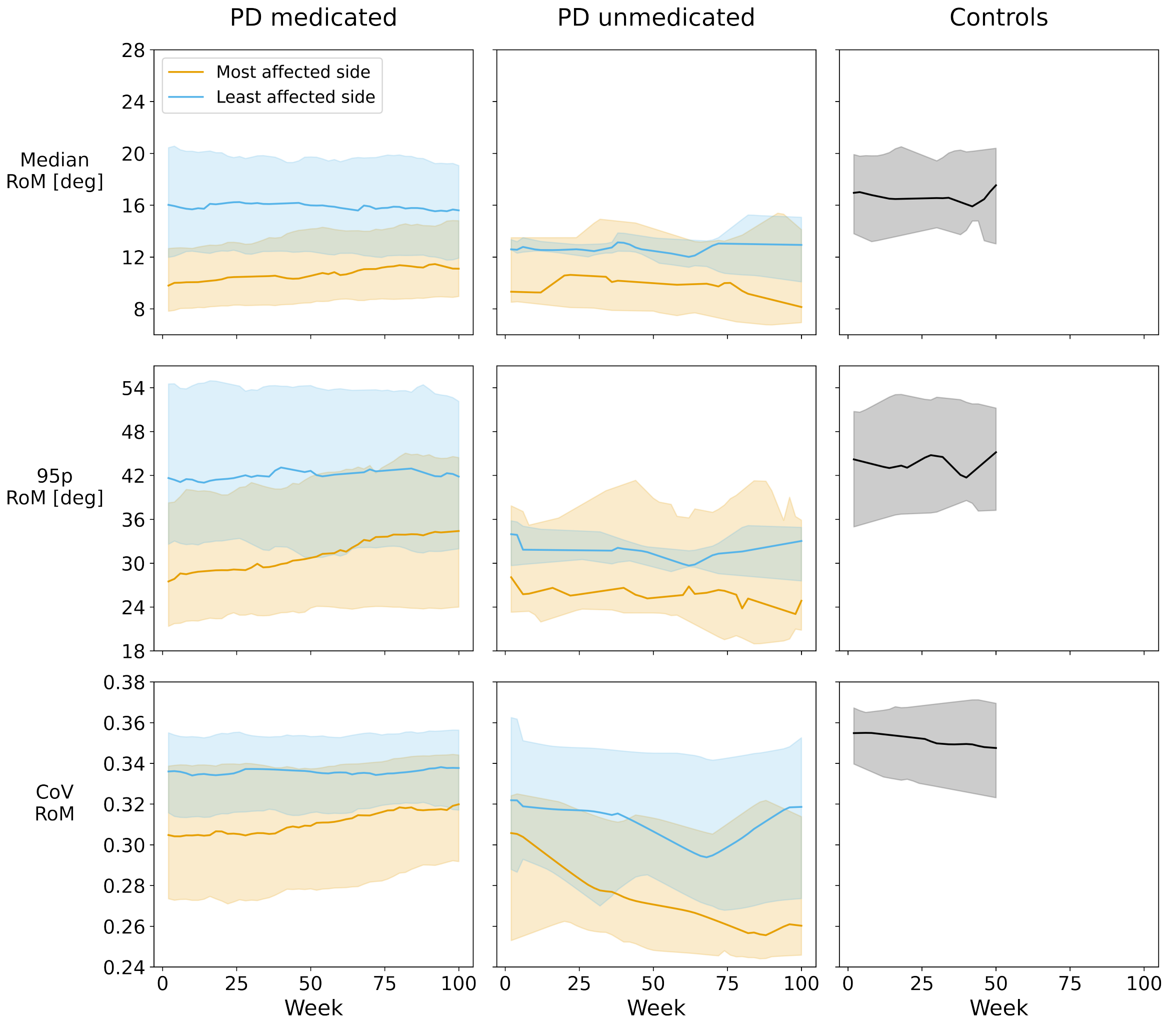


**Supplementary Figure 10: Longitudinal change in digital measures in short gait segments.** Bold lines represent the median of digital measures in short gait segments throughout the study period aggregated across participants included in the two-year SRM analysis. Shaded areas represent interquartile ranges. 95p: 95$^{th}$ percentile; CoV: coefficient of variation; deg: degrees.

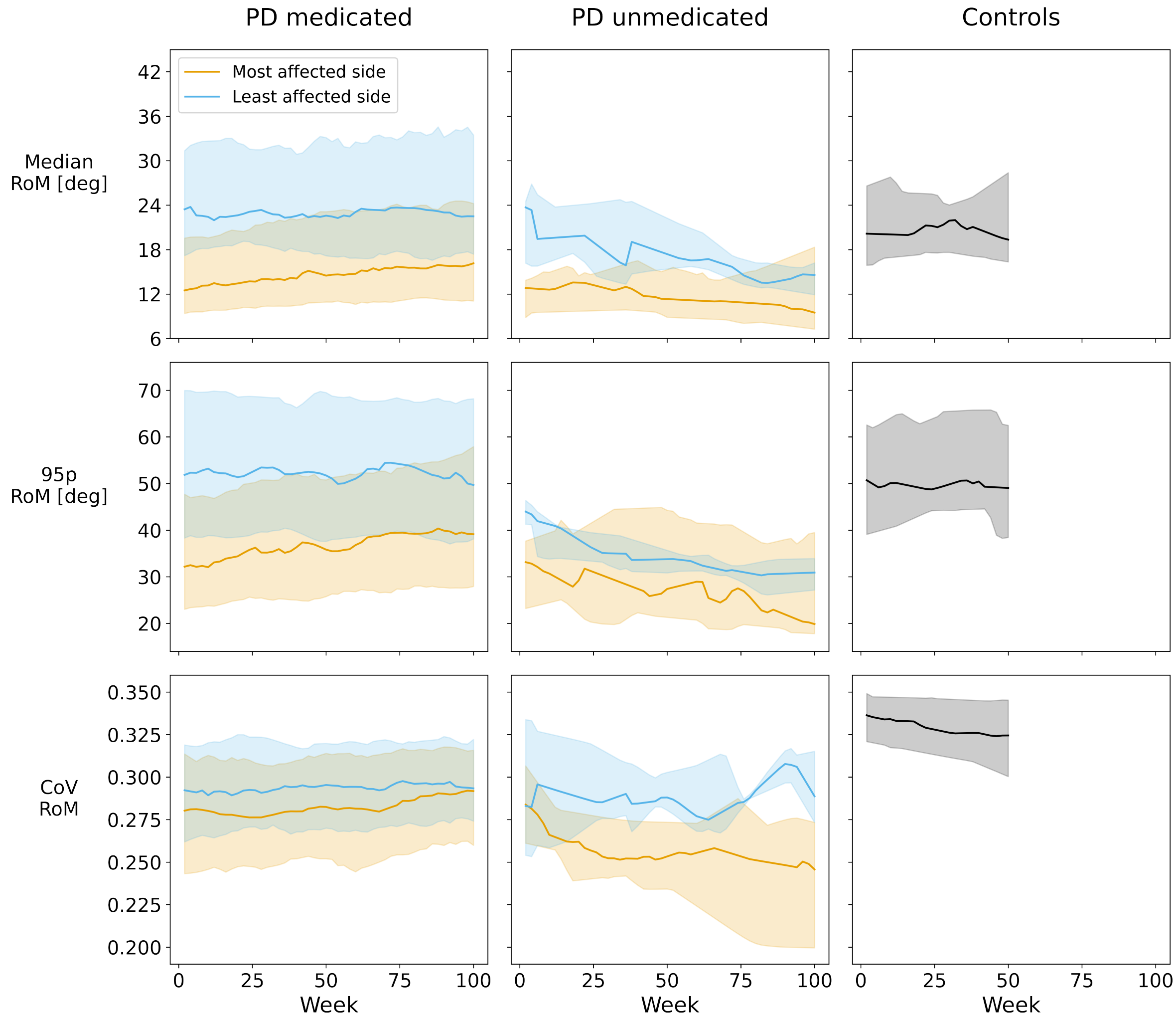


**Supplementary Figure 11: Longitudinal change in digital measures in long gait segments.** Bold lines represent the median of digital measures in long gait segments throughout the study period aggregated across participants included in the two-year SRM analysis. Shaded areas represent interquartile ranges. 95p: $95^{th}$ percentile; CoV: coefficient of variation; deg: degrees.

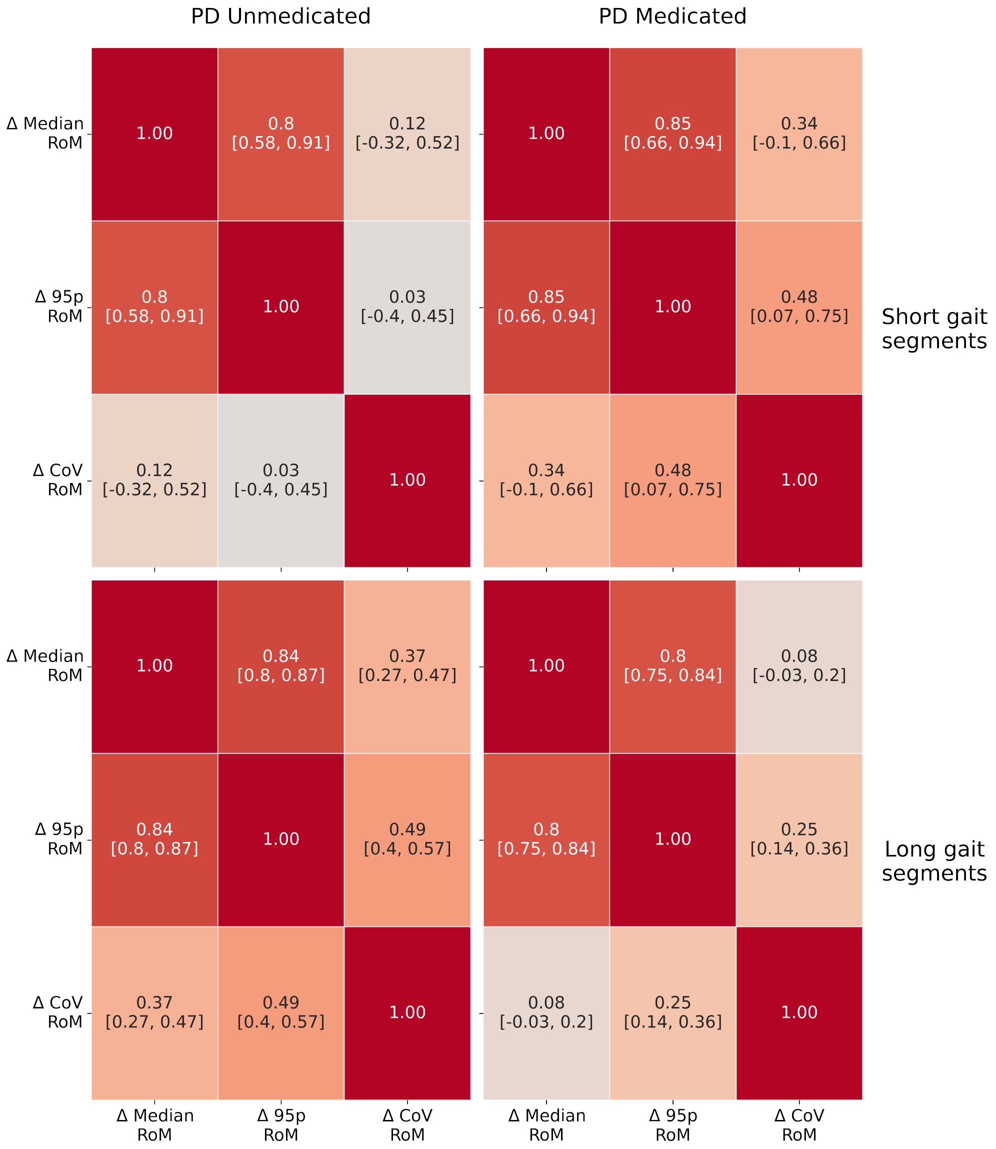


**Supplementary Figure 12: Association between change in digital measures.** Values represent Spearman rank correlations between the two-year change in digital measures. Values between squared brackets represent 95% bias-corrected and accelerated bootstrap confidence intervals. Short gait segments: $< 20$ s. Long gait segments: $\geq 20$ s. RoM: Range of motion; 95p: $95^{\text{th}}$ percentile; CoV: coefficient of variation.

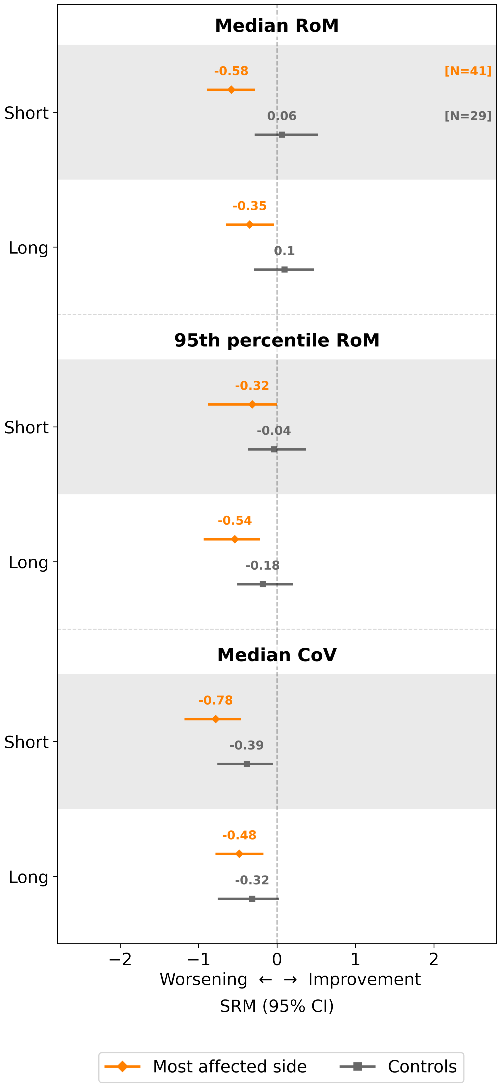


**Supplementary Figure 13: Sensitivity to one-year change of digital measures in PD compared with controls.** Values represent one-year median (95% CI) standardized response means of the digital measures for controls (gray) versus PD unmedicated wearing the watch on the most affected side (orange). RoM: range of motion; CoV: coefficient of variation. 95p: 95$^{\text{th}}$ percentile; CoV: coefficient of variation; short: gait segments $< 20$ seconds; long: gait segments $\geq 20$ seconds.

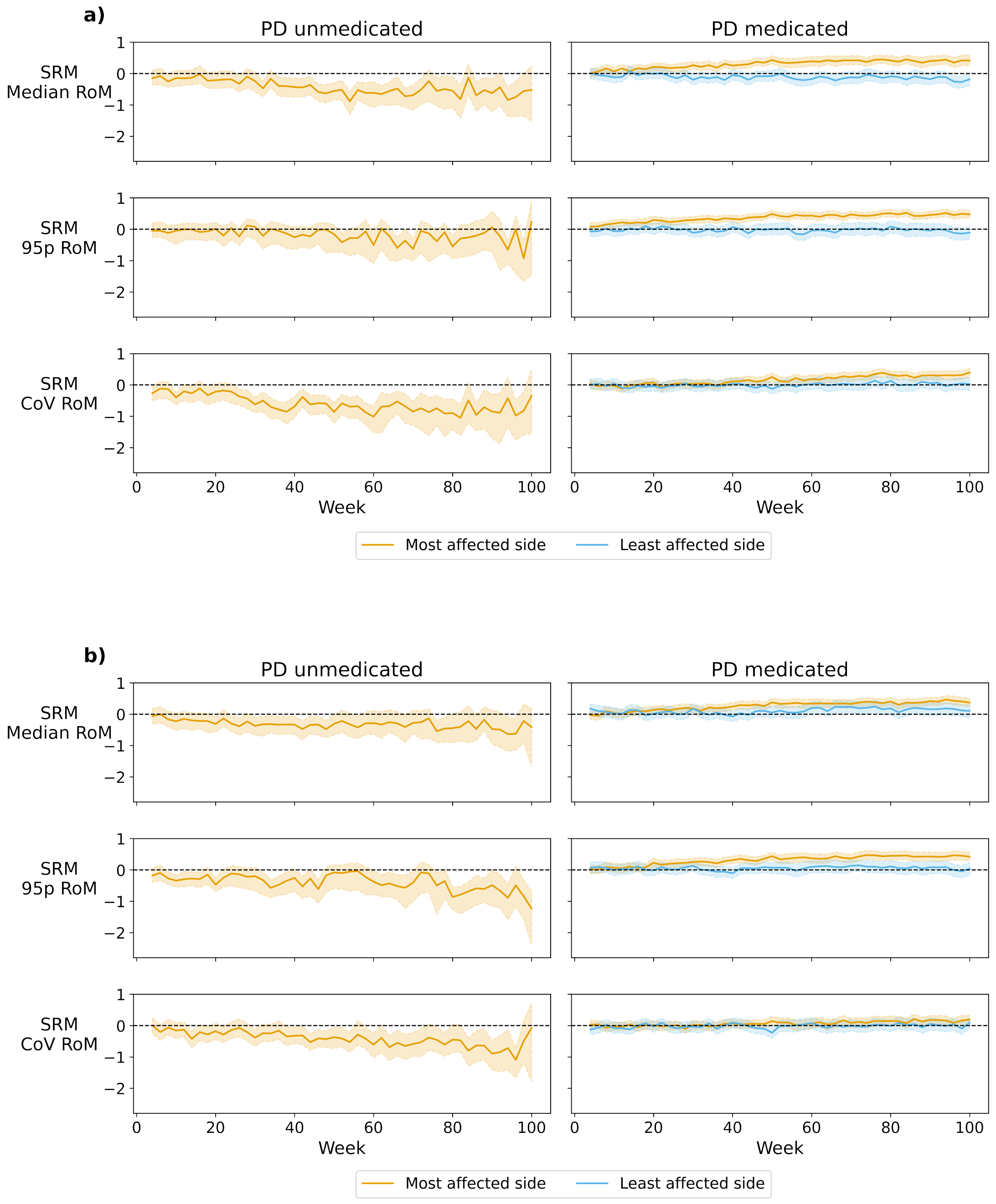


**Supplementary Figure 14: Sensitivity to change for varying study durations using biweekly measurements.** Bold lines represent standardized response means (SRMs) of biweekly measurements (instead of the trend) of the digital measures for varying study durations in **(a)** short gait segments ($< 20$ seconds) and **(b)** long gait segments ($\geq 20$ seconds). Shaded areas represent 95% bias-corrected and accelerated bootstrap confidence intervals. The number of participants varies longitudinally due to censoring and dropout. RoM: range of motion; 95p: 95$^{\text{th}}$ percentile; CoV: coefficient of variation.

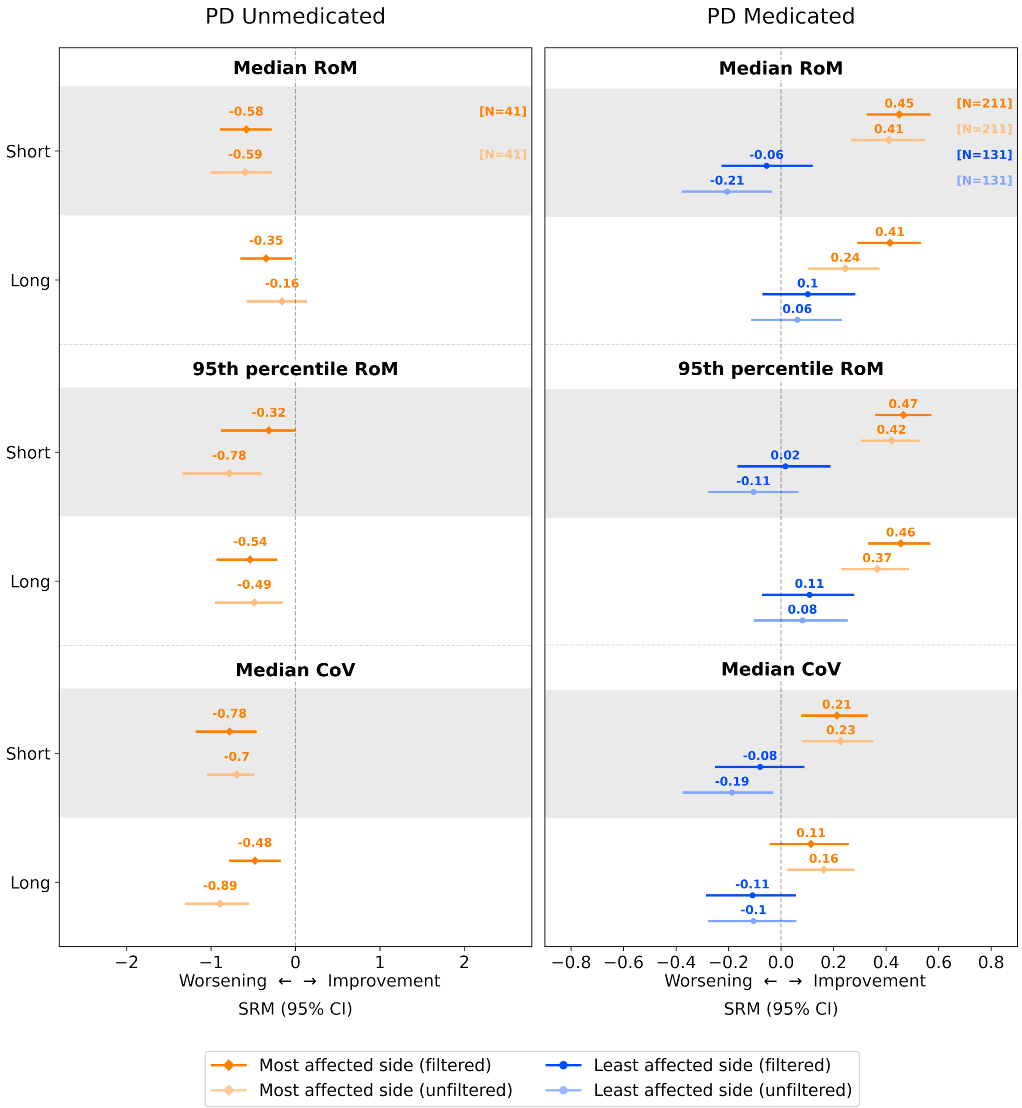


**Supplementary Figure 15: Effect of filtering on sensitivity to one-year change.** Values represent one-year standardized response means (SRMs) of digital measures. Error bars represent 95% confidence intervals (CIs). The x-axis represents worsening of arm swing for negative and improvement for positive SRM values, where worsening is defined as increasing the difference with controls. RoM: range of motion; CoV: coefficient of variation; MDS-UPDRS: Movement Disorder Society Unified Parkinson's Disease Rating Scale; short: gait segments $< 20$ seconds; long: gait segments $\geq 20$ seconds.

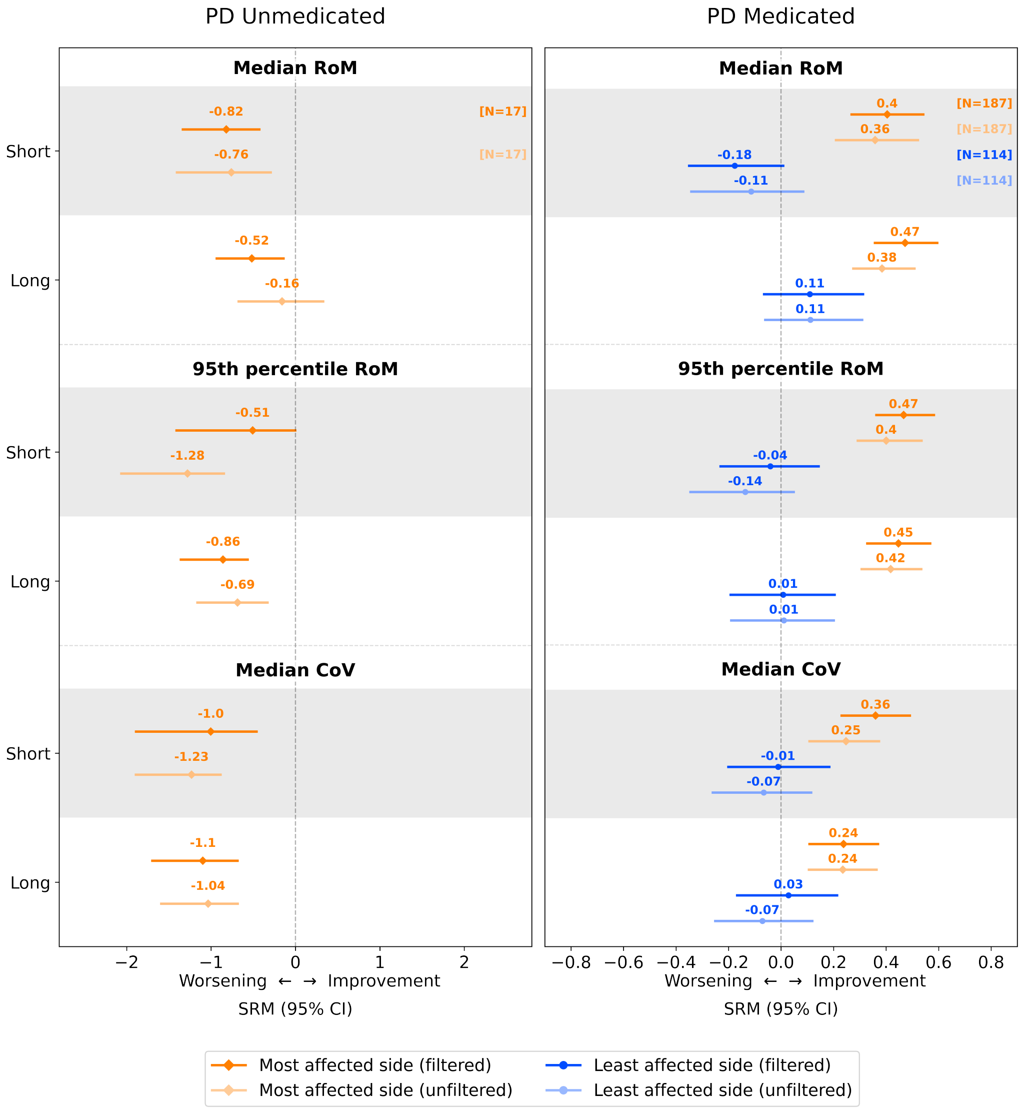


**Supplementary Figure 16: Effect of filtering on two-year change.** Values represent two-year standardized response means (SRMs) of digital measures. Error bars represent 95% confidence intervals (CIs). The x-axis represents worsening of arm swing for negative and improvement for positive SRM values, where worsening is defined as increasing the difference with controls. RoM: range of motion; CoV: coefficient of variation; MDS-UPDRS: Movement Disorder Society Unified Parkinson's Disease Rating Scale; short: gait segments $< 20$ seconds; long: gait segments $\geq 20$ seconds.

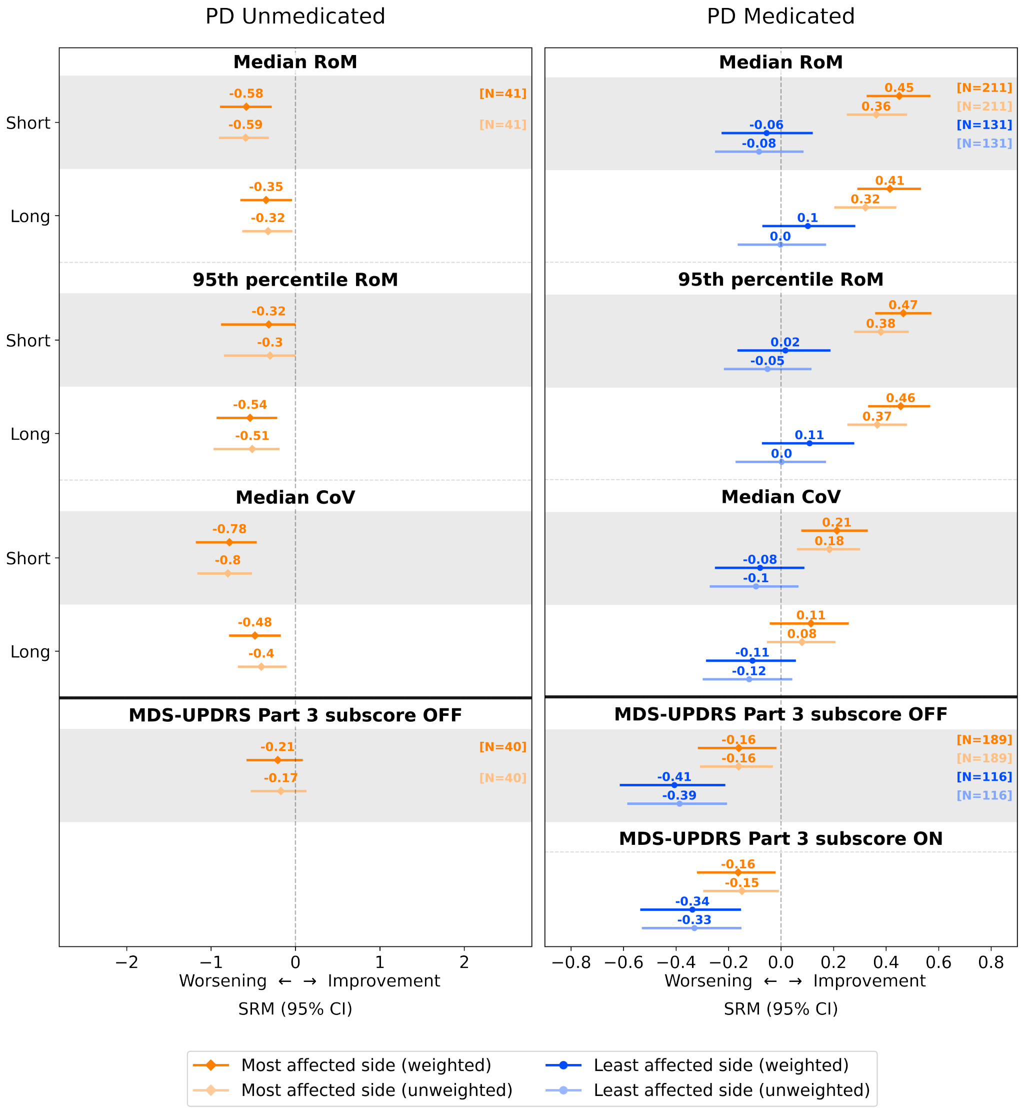


**Supplementary Figure 17: Effect of weighting on sensitivity to one-year change.** Values represent one-year standardized response means (SRMs) of digital measures in filtered gait segments and clinical measures. Error bars represent 95% bias-corrected and accelerated bootstrap confidence intervals (CIs). The sign of clinical measures is flipped such that the x-axis represents worsening of arm swing for negative and improvement for positive SRM values, where worsening is defined as increasing the difference with controls. RoM: range of motion; CoV: coefficient of variation; MDS-UPDRS: Movement Disorder Society Unified Parkinson's Disease Rating Scale; short: gait segments $< 20$ seconds; long: gait segments $\geq 20$ seconds.

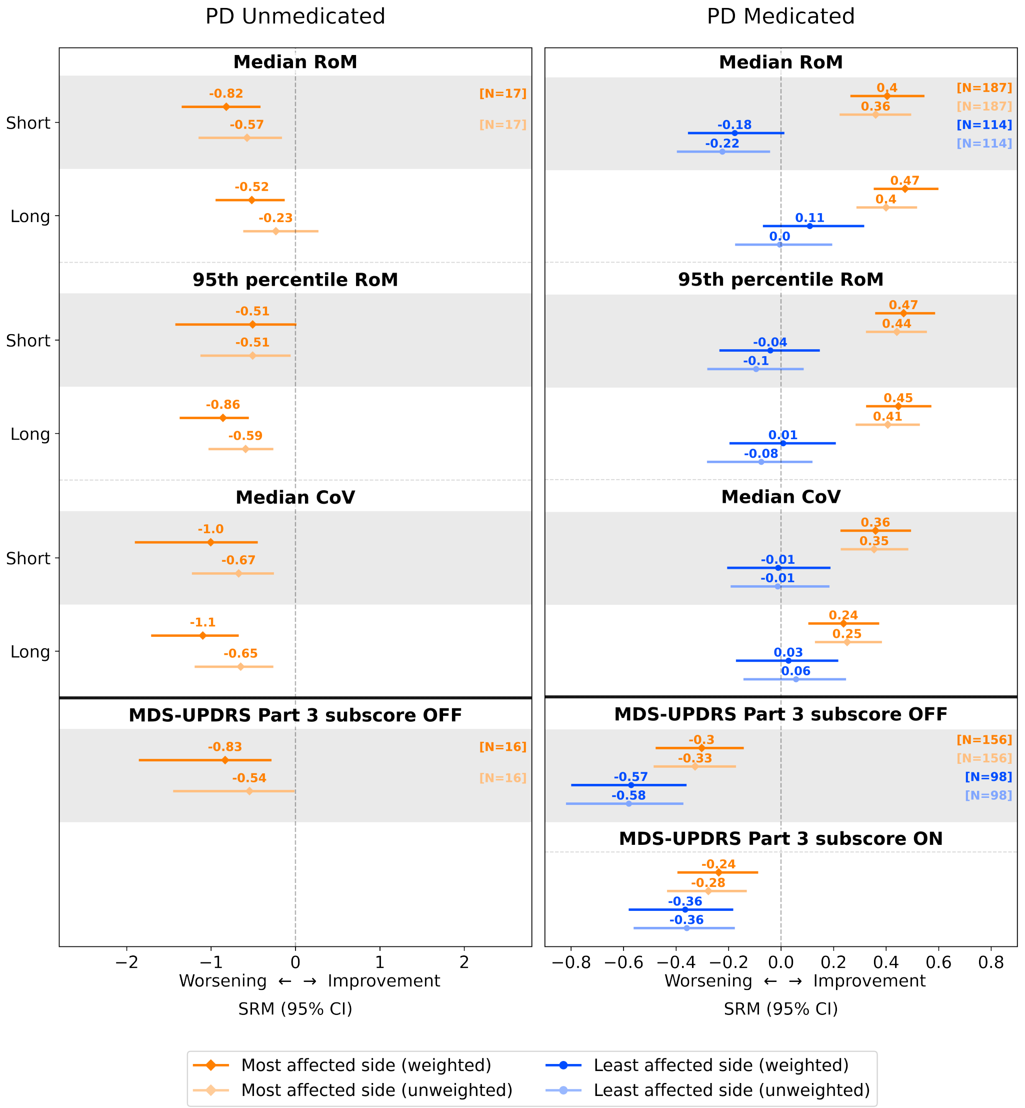


**Supplementary Figure 18: Effect of weighting on sensitivity to two-year change.** Values represent two-year standardized response means (SRMs) of digital measures in filtered gait segments and clinical measures. Error bars represent 95% bias-corrected and accelerated bootstrap confidence intervals (CIs). The sign of clinical measures is flipped such that the x-axis represents worsening of arm swing for negative and improvement for positive SRM values, where worsening is defined as increasing the difference with controls. RoM: range of motion; CoV: coefficient of variation; MDS-UPDRS: Movement Disorder Society Unified Parkinson's Disease Rating Scale; short: gait segments $< 20$ seconds; long: gait segments $\geq 20$ seconds.


1. Johansson, M. S., Korshøj, M., Schnohr, P., Marott, J. L., Prescott, E. I. B., Søgaard, K., Holtermann, A. Time spent cycling, walking, running, standing and sedentary: a cross-sectional analysis of accelerometer-data from 1670 adults in the Copenhagen City Heart Study : Physical behaviours among 1670 Copenhageners. *BMC Public Health* **19**, 1. (2019).
2. Adams, J. L., Dinesh, K., Snyder, C. W., Xiong, M., Tarolli, C. G., Sharma, S., Dorsey, E. R., Sharma, G. A real-world study of wearable sensors in Parkinson's disease. *NPJ Parkinsons Dis.* **7**, 1. (2021).
